# Multi-strain fermented milk promotes gut microbiota recovery after *Helicobacter pylori* therapy: a randomised, controlled trial

**DOI:** 10.1101/2021.01.14.21249458

**Authors:** Eric Guillemard, Marion Poirel, Florent Schäfer, Laurent Quinquis, Caroline Rossoni, Christian Keicher, Frank Wagner, Hania Szajewska, Frédéric Barbut, Muriel Derrien, Peter Malfertheiner

## Abstract

**BACKGROUND:** *Helicobacter pylori* (*Hp*) eradication therapy can alter gut microbiota, provoking gastro-intestinal (GI) symptoms that can be improved by probiotics. The effect on *Hp* patients of a Test fermented milk containing yogurt and three *Lacticaseibacillus* (*L. paracasei* CNCM I-1518, CNCM I-3689, *L. rhamnosus* CNCM I-3690) strains was assessed on antibiotic associated diarrhea (AAD) (primary aim), GI-symptoms, gut microbiota composition and metabolites. In this aim, a randomized, double-blind, controlled trial was performed in 136 adults under *Hp*-treatment (14-day amoxicillin, clarithromycin, pantoprazole), who consumed the Test or a Control product for 28 days. Feces were analysed for gut microbiota composition, short chain fatty acids (SCFA), calprotectin, and viability of ingested strains.

**RESULTS:** No effect of Test product was observed on AAD occurrence or duration, rating scores or number of days of GI symptoms. *Hp*-treatment induced a significant alteration in bacterial and fungal composition, a decrease of bacterial count and alpha-diversity, an increase of *Candida* and of calprotectin, and a decrease of SCFA concentration. Following *Hp* treatment, in the Test as compared to Control group, intra-subject beta-diversity distance from baseline was lower (p_adj_=0.02), *Escherichia-Shigella* (p_adj_=0.0082) and *Klebsiella* (p_adj_= 0.013) were significantly less abundant, and concentrations of major SCFA (p=0.035) and valerate (p = 0.045) were higher. Viable *Lp* and *Lr* strains from the Test product were mainly detected during product consumption in feces.

**CONCLUSIONS:** The study showed that 14-day *Hp* triple therapy alters gut bacterial and fungal community, their metabolites and gut inflammatory markers. Consumption of a multi-strain fermented milk can induce faster recovery of the microbiota composition and SCFA production and limit the bloom of pathobionts. (ClinicalTrials.gov, NCT02900196; First Posted : September 14, 2016; https://www.clinicaltrials.gov/ct2/show/NCT02900196).

## Introduction

Antibiotics have been reported to alter gut microbiota to variable extents, which may lead to an overgrowth of opportunistic pathogens such as *Clostridioides difficile (Cd)*, a decrease of the metabolism of primary bile acids and of non-digested carbohydrate and a reduction of short-chain fatty acids (SCFA) production ^1^. Also non-antibiotics drugs like proton pump inhibitor (PPI) can alter the gut microbiome ^2^. As a consequence, a series of gastro-intestinal (GI) symptoms including antibiotic-associated diarrhea (AAD) may result ^1 3^, and be responsible for treatment discontinuation and induction of antibiotic resistance as reported in the case of *Helicobacter pylori* (*Hp*) eradication treatment ^3^.

The recent definition of *Hp* gastritis as an infectious disease has extended the indication for therapy independent of the presence of symptoms or clinical complications ^4 5^. In regions with low clarithromycin resistance (<15%), PPI-based standard triple treatment with clarithromycin and amoxicillin (or metronidazole) is a recommended first line therapy for *Hp* eradication ^5 6^. Several studies reported that standard triple therapy can induce alteration of gut microbiota (reduced alpha-diversity and altered relative abundance of bacterial genera), that might persist for several months as measured by 16S rRNA gene sequencing ^7-11 12^. However, no study took the loss of bacterial load into account, allowing a quantitative microbiome profiling ^13^. The individual effects of amoxicillin or metronidazole in reducing the production of SCFA were reported ^14-16^ but the effect of complete *Hp* treatment has never been assessed. Notably, a decrease of the abundance of the butyrate producer *Faecalibacterium prausnitzii* ^17^ and alteration of gut microbiota composition associated with GI-symptoms including diarrhea ^17-19^ were reported in feces of patients under standard triple *Hp* therapy. However, the effects of *Hp* treatment on gut microbiota, SCFA and associated side effects have never been investigated within the same trial.

The benefit of probiotics in combination with *Hp* treatment regimen is uncertain ^3^. Meta-analyses reported that probiotics ^20 21^, either a specific strain ^22^ or multi-strain combinations ^23^ can improve *Hp* eradication rate and reduce *Hp*-treatment side effects including diarrhea. Additionally, intake of probiotics helped to reduce gut microbiota disruption induced by *H. pylori* eradication therapy or amoxicillin ^17 24^.

A dairy product containing the strain *Lacticaseibacillus paracasei* (*Lp*) CNCM I-1518 was previously shown to reduce both antibiotic-associated diarrhea (AAD) and *Cd*-associated diarrhea (CDAD) occurrence in hospitalized elderly ^25 26^ and to increase the *Hp* eradication rate in children ^27^. Recently, a 7-strain fermented milk product containing *Lp* CNCM I-1518, *Lp* CNCM I-3689, *Lacticaseibacillus rhamnosus* (*Lr*) CNCM I-3690 and yogurt strains, was shown to be safe and to induce a modest response of gut microbiota in healthy subjects ^28^.

The objective of the present trial was to assess the effect of 4-week consumption of this multi-strain product, on AAD (primary outcome), on GI symptoms, and on the gut microbiota composition and the SCFA and calprotectin production, in a population of adult dyspeptic patients treated for *Hp* eradication.

## Material and methods

### Study design

The study was monocentric, randomized, double blind, controlled, with two parallel arms (Test/Control, allocation ratio: 1-1) and an adaptive design with interim analysis. As described in **Figure 1**, the study included a screening phase, 14-days of *Hp*-eradication treatment (D0-D14), 28-days of product consumption (D0-D28) and 14-days of follow-up (D28-D42), with dietary restriction (D0-D42) (no yoghurts, probiotics in fermented dairy products or supplements). Seven visits were planned, in a clinical unit (Charité Research Organisation GmbH, Berlin, Germany): for inclusion (V1), randomization (V2-D0) and evaluation (V3-D7 to V7-D42), with blood (fasted) and stool sampling as described in **Figure 1** and **Additional file 1: methods**. Additional stools were collected at the first day and after the end of each AAD episode. The study was performed in accordance with the Declaration of Helsinki and Good Clinical Practice (International Conference on Harmonisation E6) and approved by the Ethics Committee of Charité – Campus Mitte (Application Number EA1/297/15) of the Charité – Universitätsmedizin Berlin, Germany. All volunteers provided a signed informed consent form (ICF). The trial was registered on clinicaltrials.gov (registration number: NCT02900196).

**Figure 1.**
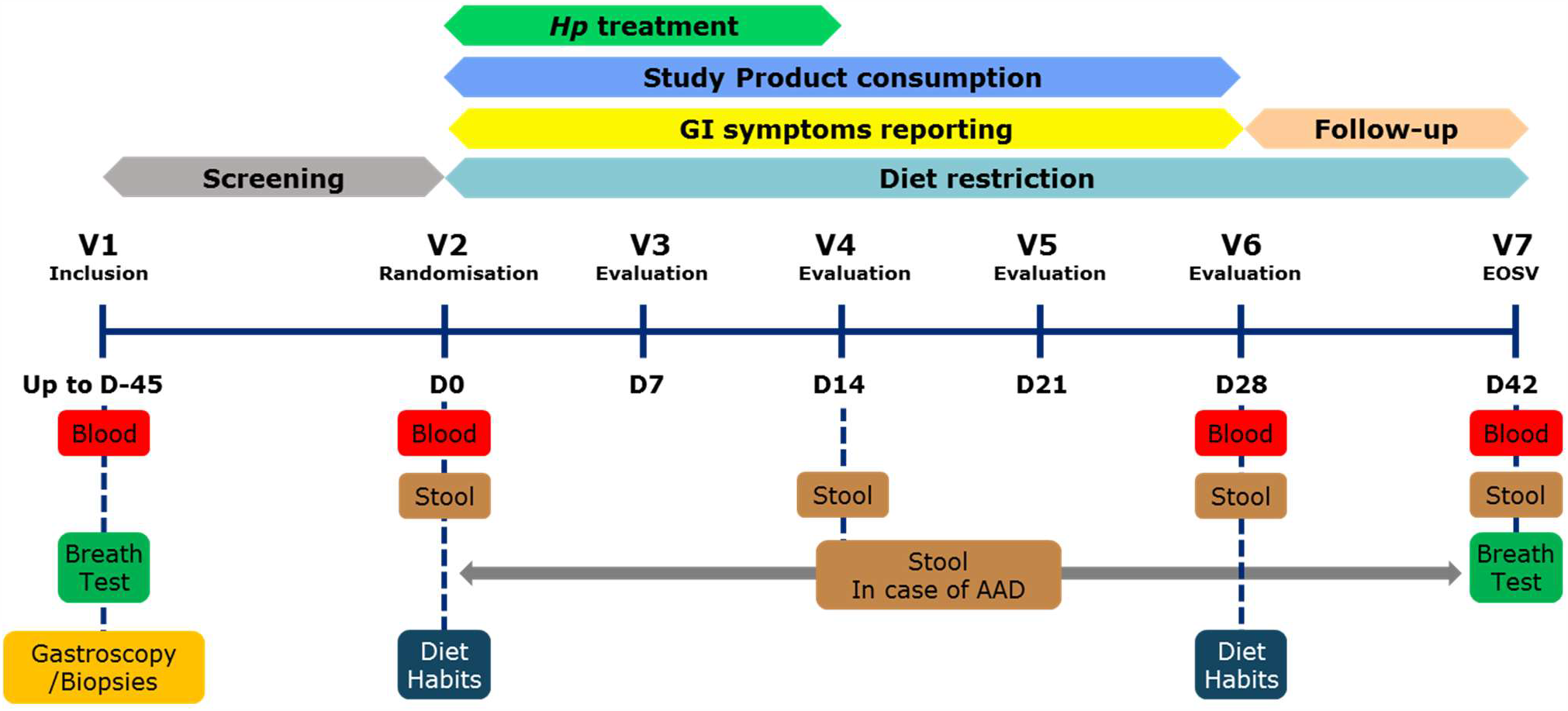
Study Design Overview.

### Subject selection

Screening lasted from September 16^th^ 2016 (first inclusion [ICF signature]) to May 31^st^ 2017 (Last inclusion), and the study experimental phase from October 07th 2016 (first randomization) to August 10^th^ 2017 (last visit).

Main inclusion criteria were: *Hp* infection, based on positive ^13^C-Urea Breath test ^29^and at least one positive Urease or *Hp*-gastritis histological test; dyspepsia with medical prescription for a *Hp*-eradication triple therapy (pantoprazole, clarithromycin and amoxicillin for 14 days); age 18 to 65 years and a body mass index (BMI) of 19 to 30 kg/m^2^.

Main exclusion criteria were: past *Hp* eradication treatment; alarm feature (bleeding, anemia, unexplained weight loss, dysphagia, odynophagia, recurrent vomiting or previous gastrointestinal malignancies); benign peptic ulcer, pre-malignant or malignant lesion; diarrhea within the preceding 4-weeks; severe evolutive, chronic or past pathology or infection of the GI-tract or surgery in the last 3 months; liver or renal diseases (based on blood transaminase and creatinine); antibiotics, intestinal antiseptic treatment during the previous 2 months or chronic use of laxatives or anti-diarrheal; H2-receptor antagonists or PPI treatment in the last 2 weeks or food allergy. Decision tree applied for screening to select *Hp* infected dyspeptic subjects without alarm symptoms or specific gastric pathologies is presented in the **Additional file 2: Figure S1**. All criteria including laboratory results from the screening period were checked again at randomization. Eligibility criteria are detailed in the **Additional file 1: Methods**.

### Product intervention and Hp eradication treatment

Test product was a fermented milk containing *Lp* CNCM I-1518, *Lp* CNCM I-3689 and *Lr* CNCM I-3690 strains and four yogurt strains. Strain counts are provided in **Additional file 3: Table S1**. Control product was an acidified milk, depleted in lactose, containing phosphoric acid and Carboxy Methyl Cellulose. Both products were manufactured by Danone Research, France, and were similar in sweetness, flavour (multi-fruit), texture, colour, packaging and nutritional content (isocaloric) to ensure the double-blinding for participants and study personnel including the outcome assessors. Subjects ingested two bottles (100 g/bottle) of Test or Control product per day (one at breakfast, one at dinner), for 28 days.

For *Hp*-eradication, subjects were treated by a triple therapy (ZacPac®, Takeda, Singen, Germay) including a PPI (pantoprazole 40mg), and two antibiotics (clarithromycin 500mg and amoxicillin 1000mg), twice daily, for 14 days.

### Outcomes

The primary outcome was the occurrence of AAD (number of subjects presenting at least one AAD episode) during the 28-day product consumption period. AAD was defined as three or more stools at 5 to 7 in Bristol Stool Scale [BSS]) per day for at least 3 consecutive days (definition 1 for main analysis) or for at least 1 day (definition 2 as secondary analysis of the primary outcome). Secondary outcomes included: AAD occurrence with alternative definitions (Two or more BSS 5-7 stools per day for at least 1 or 3 days; Three or more BSS 5-7 stools per day for at least 2 days or more than 6 BSS 5-7 stools for at least one day); AAD duration; time to event of AAD; occurrence, duration or time to event of CDAD (AAD with a positive test for *Cd*, method is provided in **Additional file 1: Methods**); cumulative number of days with GI-symptoms (diarrhea, abdominal pain, bloating, nausea or vomiting); score of Gastrointestinal Symptom Rating Scale [GSRS] questionnaire (based on a 7-graded Likert scale). Biological parameters measured in feces included microbiota composition and quantification of short and branched chain fatty acids (SCFA), further categorized as major (acetate, propionate, butyrate) and minor (valerate, caproate, isobutyrate, isovalerate), of calprotectin, and of total and viable *Lp* and *Lr* strains from Test product. SCFA were also quantified in blood. Monitored safety parameters were vital signs, anthropometry, blood analyses and adverse events (AE). Details for biological and safety parameter analyses are provided below or in **Additional file 1: methods**.

### Procedure

Subjects reported their physical activity and smoking habits (V2), dietary habits (V2 and V6), and alcohol consumption (all visits). In an e-diary, they reported daily their compliance (to *Hp* treatment, product consumption and dietary restrictions), AE, medication or supplement intake, all passages of stool and consistency (BSS), GI symptoms, and weekly reported their GSRS scores. ^13^C-urea breath test was done at V1 for *Hp* infection diagnosis and V7 for eradication measure. The study was performed in accordance with the protocol with no change during the study. Subject follow-up and detailed procedures are provided in **Additional file 1: methods**.

### Sample size

Sample size was computed based on an expected reduction of 50% (RR = 0.5) of the number of subjects experiencing AAD, a 15% AAD occurrence in the Control group and a 10% drop-out rate ^30-32^. A total of 295 randomized subjects was initially planned. After an interim analysis, the study was stopped due to futility of clinical outcomes and the final sample size was 136 randomized subjects. Interim analysis process is described in **Additional file 1: methods**.

### Randomization

A permuted block randomization (1:1 ratio) was made using an Interactive Web Response System. The randomization list was generated by an external statistician and kept confidential at the Sponsor’s premise to ensure allocation concealment. Subjects were automatically assigned to the Test or Control group. The allocation was blinded to investigator and sponsor. An independent access was provided to the technician responsible for product preparation.

### Statistical analysis

#### Clinical outcomes

Analyses were performed using SAS 9.4 software on the Full Analysis Set (FAS) population. The primary and secondary outcomes were described by product group and by visit. Due to the low number of AAD observed, no statistical tests were performed on related outcomes. A descriptive analysis of the number of days with GI symptoms and of GSRS scores (total, individual and by symptom dimension scores and their change from V2) was performed.

#### SCFA, calprotectin, quantification of Test product strains in feces

The effect of product (Test, Control) was assessed on SCFA, calprotectin and their changes from V2, V4 and V6, using a repeated linear mixed model including the values at baseline, the visit and a Group-Visit interaction as covariates. For Test product strains analysis, the effect of different time points was assessed using a Friedman (for total and viable cell count) and a Wilcoxon test (for viability loss and rate). More details are provided in **Additional file 1: methods**.

#### Gut microbiota

Statistical analyses were performed, and graphs were plotted with R software (version 3.6.0). Total bacterial count (as log_10_-transformed), alpha diversity, Shannon index ITS/16S ratio were analyzed as change from baseline using linear mixed model (nlme 3.1-140) as described above or using Mann-Whitney or Wilcoxon signed rank test when normality was rejected. Beta-diversity was analyzed using PERMANOVA (vegan 2.5-6) and pairwise group mean dispersions using Tukey HSD. Quantitative Microbiome Profiling (QMP) was done as described by Vandeputte et al. ^13^. Differential analyses were performed with DESeq2 (version 1.24.0) on 16S based QMP and ITS datasets. Fold changes were evaluated with the Wald test (FDR adj. p < 0.1). P-values were adjusted for multiple testing with the Benjamini-Hochberg procedure (p_adj_). Methods are detailed in the **Additional file 1: methods**.

## Results

### Subject enrollment, population at baseline and compliance

Of the 1012 subjects included and screened, 136 subjects (FAS population) were randomized (68 in each group) (**Additional file 4: Figure S2**). One subject withdrew from the Test group before V3 due to *Hp* treatment intolerance but was included in the FAS.

Subject characteristics at baseline (**Table 1**) were well-balanced between groups for age, sex, BMI, physical activity (IPAQ) and medical or surgical history. Proportions of alcohol consumers and smokers were slightly higher in the Test group whereas smokers in the Control group consumed a higher number of cigarettes per week. A higher proportion of subjects with concomitant medication, mainly estrogens/progestogens and anti-inflammatory medication, was reported in the Test group. Reporting of dietary habits (FFQ) showed no difference between groups in nutrient intake at V2 and V6. The observed differences in baseline characteristics were considered to have no expected impact on study product effect evaluation.

**Table 1:** Subject characteristics at baseline.

|  |  | **Test (N=68)** | **Control (N=68)** |
| --- | --- | --- | --- |
| **Age (years)** | Mean (SD) | 42.1 (10.1) | 42.6 (11.3) |
|  | Min ; Max | 26 ; 65 | 23 ; 64 |
| **Sex, n (%)** | Male | 34 (50.0) | 35 (51.5) |
|  | Female | 34 (50.0) | 33 (48.5) |
| **BMI (kg/m<sup>2</sup>)** | Mean (SD) | 24.8 (2.9) | 25.0 (2.6) |
|  | Min ; Max | 19.6 ; 30.0 | 19.7 ; 29.5 |
| **Alcohol Consumer, n (%)** | No | 17 (25.0) | 24 (35.3) |
|  | Yes | 51 (75.0) | 44 (64.7) |
| **Alcohol units/week<sup>1</sup>** | Mean (SD) | 2.2 (1.8) | 2.3 (1.4) |
|  | Min ; Max | 1 ; 8 | 1 ; 7 |
| **Smoking status, n (%)** | Current smoker | 19 (27.9) | 14 (20.6) |
|  | Ex-smoker | 20 (29.4) | 20 (29.4) |
|  | Never a smoker | 29 (42.6) | 34 (50.0) |
| **Number of cigarettes/week<sup>2</sup>** | Mean (SD) | 41.2 (39.9) | 58.1 (40.8) |
|  | Min ; Max | 1 ; 140 | 1 ; 105 |
| **Physical activity IPAQ, n (%)** | Low | 8 (11.8) | 8 (11.8) |
|  | Moderate | 24 (35.3) | 22 (32.4) |
|  | High | 36 (52.9) | 38 (55.9) |
| **Medical/Surgical history, n (%)** |  | 49 (72.1) | 47 (69.1) |
| **Concomitant medication or nutritional supplements, n (%)** |  | 38 (55.9) | 27 (39.7) |
| **Product restricted during the study, n (%)** |  | 0 | 0 |

<sup>1</sup> : In alcohol consumer
<sup>2</sup> : in current smokers
IPAQ = international physical activity questionnaire,

Subject compliance (mean [SD]) was high in both Test and Control groups for study product consumption (99.2 [3.3] % and 99.7 [1.0] %) and for *Hp* treatment (97.2 [12.2] % and 98.4 [7.7] %). Major deviations to the protocol were reported in only 14 (10.3%) subjects (7 in each group), mainly due to low compliance to *Hp* treatment or study products consumption. A small amount of missing data was reported for clinical parameters (<10%).

### Clinical outcomes

At V7, *Hp* eradication was successful in 83.8% and 88.2% of subjects in Test and Control groups respectively. As primary outcome, the occurrence of AAD was much lower than expected since only 3 and 8 subjects reported an episode as per definition 1 and 2, respectively, in the FAS population (**Table 2**). Due to the limited number of episodes, no statistical test could be applied for the main analysis of the primary outcome or for any other clinical outcomes consistent with the planned strategy to handle multiple testing. All clinical criteria were analyzed based on descriptive statistics. The limited number of events did not allow any conclusion on the product efficacy on AAD occurrence (**Table 2**). No CDAD was reported and only two *Cd* positive subjects were found in the Test group, likely as a consequence of *Hp*-treatment (**Table 2**). No relevant difference between groups was observed for other clinical outcomes, as described in additional files **(Additional file 5: Results, Additional file 6: Table S2, Additional file 7: Figure S3**) including occurrence of AAD with alternative definition, time to first event, AAD duration, number of days with GI symptoms or GSRS scores.

**Table 2.**
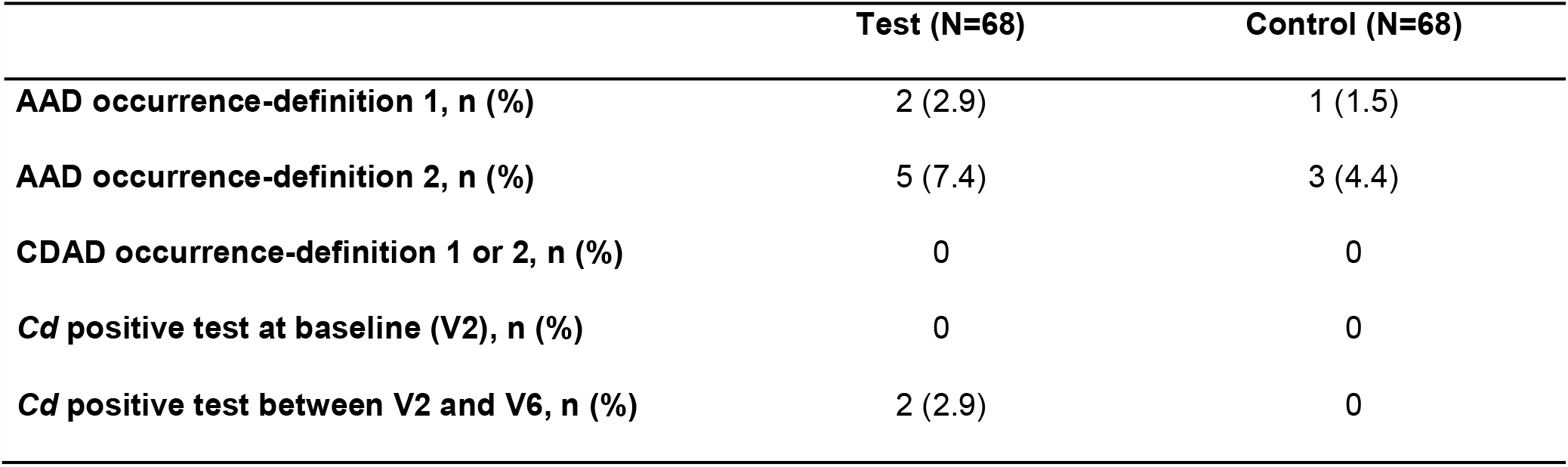
Occurrence of AAD and CDAD. Occurrence of AAD and CDAD during the 28-day product consumption period. AAD Definitions: Three or more BSS 5-7 stools/per day for at least 3 days (Definition 1) or at least 1 day (Definitions 2).

### Test product strains viability in feces

The viability of the three strains *Lp* CNCM I-3689, *Lr* CNCM I-3690 and *Lp* CNCM I-1518 was assessed in the feces of 48 subjects (25 in Test, 23 in Control groups) (**Additional file 8: Figure S4**). At V4, total cell (live and dead) counts reached 8.1 to 8.3 log_10_ cells/g feces depending on the strain with a viability rate of 2.2% (*Lr* CNCM I-3690), 2.3% (*Lp* CNCM I-3689) and 9.4% (*Lp* CNCM I-1518). At V6, no significant change was observed for either the total or viable cell counts. At V7, two weeks after the end of the last Test product intake, the *Lr* CNCM I-3690 and *Lp* CNCM I-3689 strains were found viable in 2 and 1 subjects respectively. Additional data are provided in **Additional file 5: Results**.

### Global gut microbiota response to Hp treatment and product intervention

Gut microbiota was analyzed in 135 subjects (67 in Test and 68 in Control group) using flow cytometry and 16S rRNA gene sequencing. In both groups, there was a significant loss of total bacteria at V4, after 2 weeks of products intake and concomitant *Hp*-treatment (-0.4 log10, p_adj_ = 8.66^E-24^) (Figure 2A). Bacteria count then gradually increased until V7, 28 days following cessation of *Hp* treatment, to a level lower than that of baseline (-0.13 log10, p_adj_ = 5.34^E-05^). No global difference between Test and Control groups was observed across time points (p = 0.64) **(Figure 2A)**. The same kinetics was observed for alpha-diversity indexes, assessed by Shannon and reciprocal Simpson, which decreased at V4 (resp. p_adj_ = 3.19^E-28^, p_adj_ = 3.08^E-32^), and gradually increased without reaching baseline level at V7 (resp. p_adj_ = 1.25^E-07^, p_adj_ = 1.39^E-10^), with no difference observed between groups (resp. p_adj_ = 0.74, p_adj_ = 0.49) **(Figure 2 B and C)**. Beta-diversity, assessed by Principal Coordinate Analyses (PCoA) for Bray-Curtis dissimilarity, revealed a shift in the global microbiota composition at V4 as compared to baseline (PERMANOVA, p_adj_=1.25^E-3^ **(Figure 2D)** and a gradual but incomplete recovery at V7 (PERMANOVA, p_adj_ =1.25^E-3^). This was also observed with UniFrac distances **(Additional file 9: Figure S5)**. We observed higher variability in beta-diversity following *Hp* treatment than at baseline (Tukey HSD, p_adj_ =3.31^E-10^).

**Figure 2.**
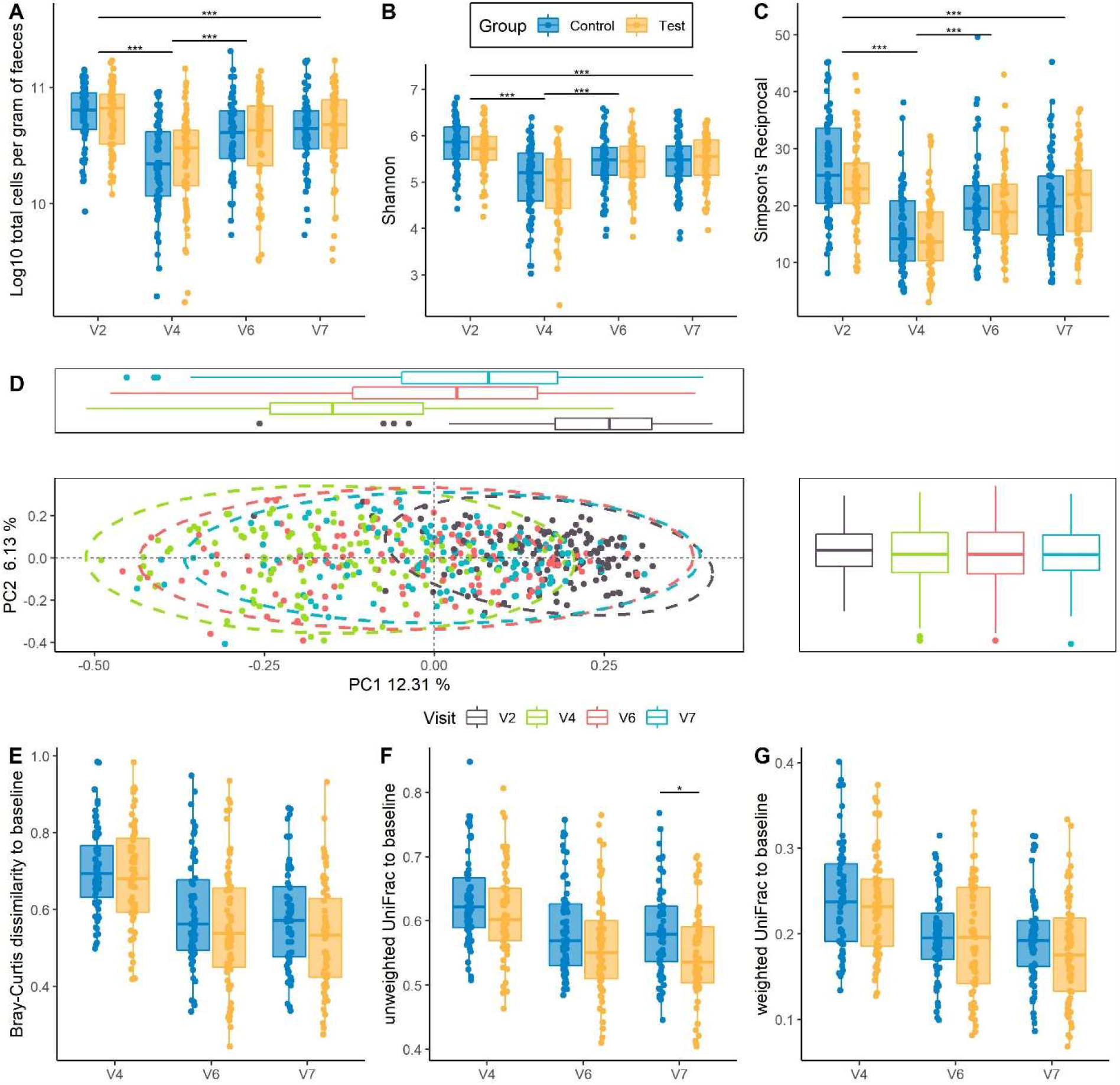
Global gut microbiota response to *Hp* treatment and product intervention. (A) Total bacterial count / g fecal samples assessed by flow cytometry, with a log10 transformation. (B) Alpha-diversity assessed by Shannon index (C) Alpha-diversity assessed by Simpson’s Reciprocal. (D) Principal Coordinate analysis (PcoA) based on Bray-Curtis dissimilarity. Samples were collected before (V2) and after *Hp* treatment (V4), 14 days (V6) and 28 days (V7) following cessation of *Hp* treatment. * p <0.05; ** p < 0.01; *** p < 0.001 according to linear mixed model. (E-G) Intra-subject distance to baseline of each subject in Test and Control groups across the study (E). Bray-Curtis dissimilarity (F) Weighted UniFrac distance (G) Unweighted UniFrac distance. * p <0.05; ** p < 0.01; *** p < 0.001 according to Mann-Whitney test.

Intra-subject distance to baseline was computed for the three beta-diversity metrics between baseline and each of the following time points. A gradual decrease of intra-subject distance in both groups following cessation of *Hp* treatment was observed with no difference between groups based on Bray-Curtis dissimilarity (Mann-Whitney test, p_adj_ = 0.12 at V7) and Weighted UniFrac **(Figure 2 E and F)**. Unweighted UniFrac intra-subject distance tended to be lower in Test-group at V4 and V6 (Mann-Whitney test, p_adj_ = 0.07 for both) and was significantly lower at V7 (Mann-Whitney test, p_adj_ = 0.02) **(Figure 2G)**. Results from alpha and beta-diversity measured by quantitative microbiome profiling (QMP) were consistent with that of relative microbiome profiling (**Additional file 10: Figure S6)**. In addition, lower intra-subject distance was observed in the Test group at V7 for Bray-Curtis dissimilarity by QMP (Mann-Whitney test, p_adj_ = 0.03) **(Additional file 10: Figure S6)**.

### Taxonomical analysis by Quantitative Microbiome Profiling

DESeq2 analysis showed that 14-day *Hp* treatment induced differential quantitative abundance of taxonomically diverse genera in the Control group (see **Additional file 11: Table S3** for Fold change and adjusted p values). Genera with depleted abundance following *Hp* treatment included members of Actinobacteria (*Bifidobacterium, Collinsella, Olsenella, Slackia*) **(Figure 3A)**, while *Eggerthella, Bacteroides*, and some members from Enterobacteriaceae (*Escherichia-Shigella, Klebsiella*) were enriched. The short-term response was variable between genera **(Figure 3B)**. Genera whose abundance returned to baseline level included *Succinivibrio, Coprococcus, Dialister* **(Figure 3B)**. Bacterial genera whose response differed between Test and Control following *Hp* treatment included *Slackia, Desulfovibrio* that were enriched in Test, while *Fusobacterium, Coprobacter, Escherichia-Shigella* were depleted (**Figure 3C)**.). Given the difference between groups observed based on beta-diversity, on unweighted UniFrac and Bray-Curtis dissimilarity at V7, we further identified genera whose abundance recovered differently from *Hp* treatment between groups **(Figure 3C** and **Additional file 11: Table S3)**. We observed that fewer genera were differently abundant between baseline and V7 in the Test (93 genera) compared to the Control group (102 genera) **(Figure 3C** and **Additional file 11: Table S3)**. Amongst them, *Escherichia-Shigella* and *Klebsiella* were significantly enriched while *Methanobrevibacter, Butyrivibrio* were significantly depleted, in the Control but not in the Test group. In contrast, the abundance of *Roseburia, Succinivibrio*, and *Dialister* was lower at V7 in the Test group but not in the Control (**Figure 3C)**.

**Figure 3.**
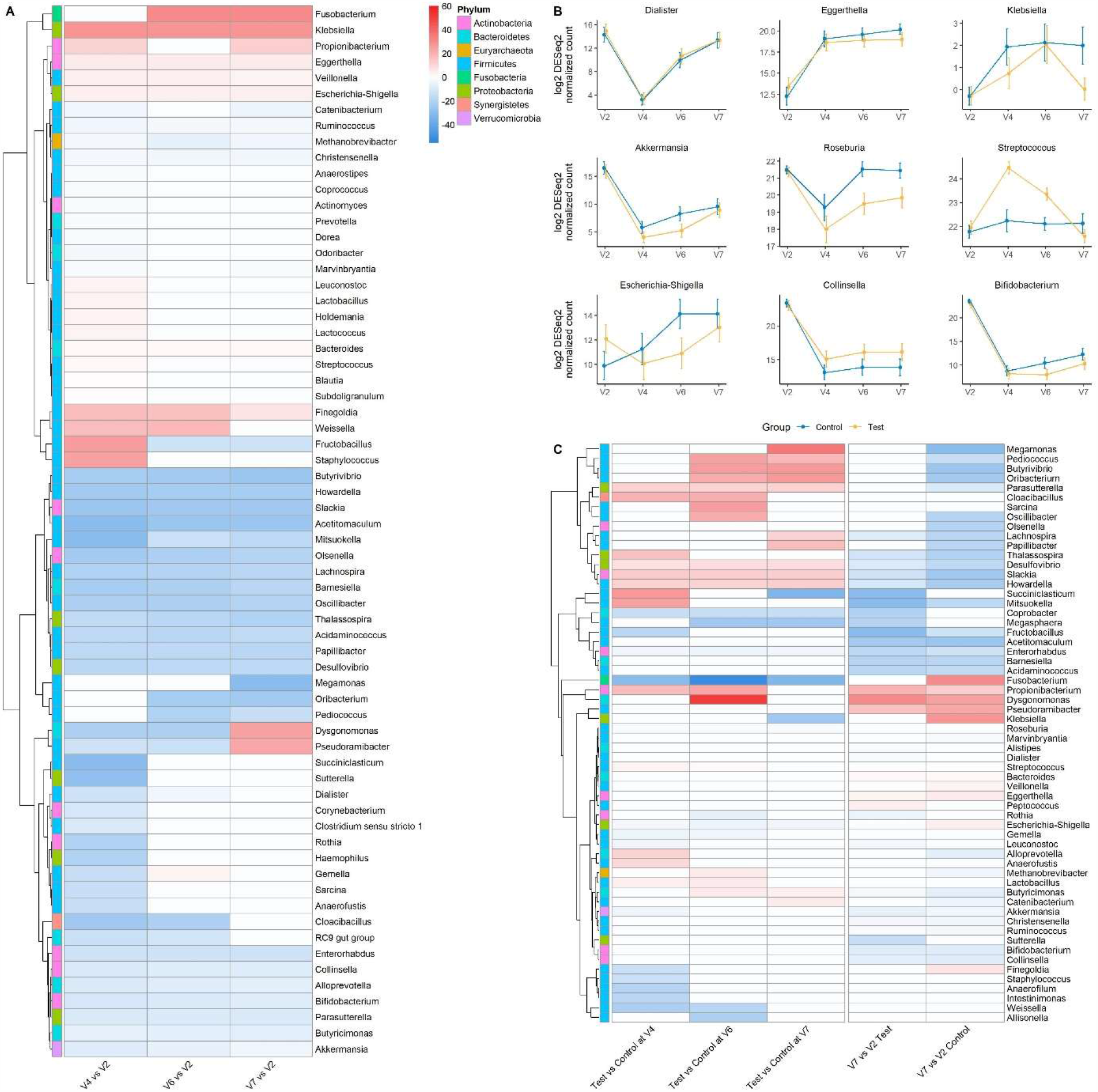
Genus-level differential analysis based on Quantitative Microbiome Profiling. (A) Heatmap of differentially abundant genera between each visit and baseline (V2). (B) Quantitative abundance of selected genera displaying different dynamic patterns in response to *Hp* treatment along the study, as mean ± 95% CI. C. Heatmap of differentially abundant genera between Test and Control groups at each visit, and between V7 and baseline (V2) within groups. For both heatmaps, only assigned genera were selected based on DESeq2 normalized counts >10.000), (FDR adj. p < 0.1, DESeq2-based Wald test). Red indicates higher fold-change and blue lower fold-change with regard to the reference (V2 or Control). Non-significant fold changes were set to zero for heatmap display.

### Gut mycobiota response to *Hp* treatment and product intervention

We monitored fungal dynamics across the study. While there was no change in ITS Shannon index in response to *Hp* treatment **(Figure 4A)**, we observed a transient increase in the fungi to bacteria (ITS/16S) Shannon ratio at V4 (p_adj_ = 5.79^E-03^) which then reverted to baseline at V7 (p_adj_ = 0.864), with a trend for a lower ITS/16S ratio (p = 0.08) in the Test group (**Figure 4B)**. The abundance of *Candida* was transiently enriched following 14-day *Hp* treatment (_adj_= 1.81^E-25^), followed by a recovery to baseline level (V2 to V7 p_adj_ = 0.467), with no difference between groups (**Figure 4C)**. Additional data are provided in **Additional file 5: Results**.

**Figure 4.**
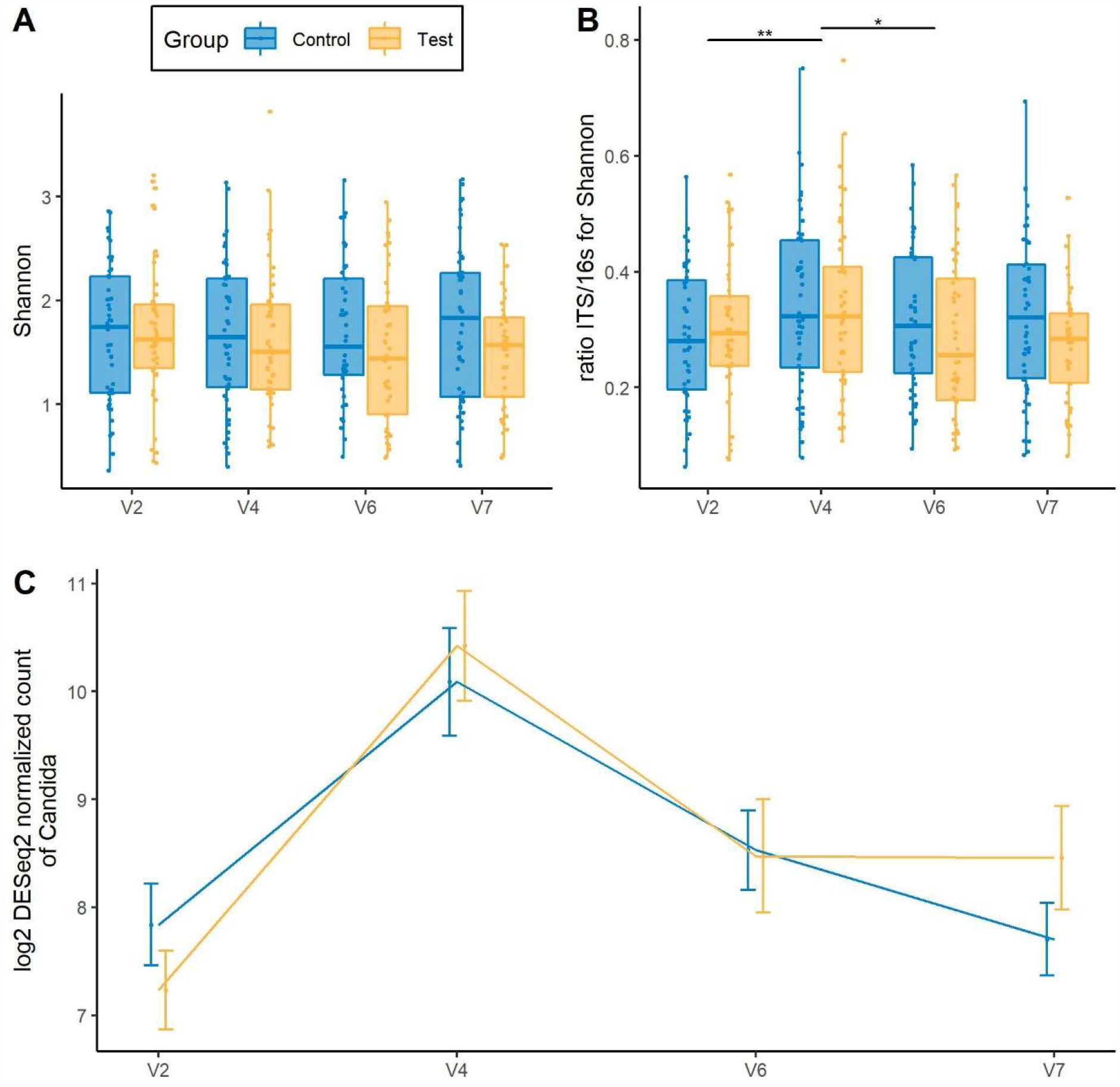
Global gut mycobiota response to *Hp* treatment and product intervention. (A) Alpha-diversity assessed by Shannon index (B) Shannon index ratio ITS/16S based C. Abundance of *Candida*

### SCFA and Calprotectin response to H pylori treatment and product intervention

Major and minor SCFA were quantified in dry feces from 61 subjects (31 and 30 subjects in Test and Control group respectively). In both groups, SCFA concentration significantly decreased at the end of *Hp*-treatment (V4) as compared to baseline (V2) for major SCFA (p = 0.0167 in Test, p = 0.0040 in Control) (**Figure 5A, B**), minor SCFA (p <0.0001 in both groups) (**Figure 5C, D**), and similarly for each SCFA type (p <0.05) (**Additional file 12: Figure S8**). From V4 to V6, at the end of product consumption, SCFA concentrations globally further decreased in the Control group and systematically increased in the Test group for all SCFA types and categories, albeit with no statistical significance within each group (**Figure 5** and **Additional file 12: Figure S8**). Evolution in both groups resulted in SCFA concentrations at V6 that were all higher and closer to baseline values in the Test as compared to the Control group. A statistically significant difference between groups was observed in the change from V4 to V6 for major SCFA resulting in concentrations (Mean (SD)) of 492.44 (247.3) and 366.7 (178.6) µmol/g in Test and Control groups respectively, corresponding to a difference of changes (Mean [95% CI]) of 120.83 [8.21;233.45] µmol/g (Student’s test, p=0.035) (**Figure 5A, B**). For valerate, a significant difference of change between groups (Mean [95% CI]) of 2.60 [0.06;5.15] µmol/g (Student’s test, p = 0.045) was also observed from V2 to V6 (**Additional file 12: Figure S8C, S8D**), corresponding to 32% less decrease of valerate concentration in the Test group. From V6 to V7, 2 weeks after product consumption ceased, the results showed a trend of stabilization (for major SCFA) or an increase (for minor SCFA) of the concentrations in the Test group and a systematic increase of both in the Control group. This resulted in a similar non statistically different levels of all SCFA concentrations between groups at V7. As compared to baseline, concentrations at V7 were mostly lower, with a significant statistical difference for major SCFA in the Test group and for minor SCFA in both groups (**Figure 5** and **Additional file 12: Figure S8**). Serum contents of SCFA showed no difference between groups at any timepoint between V2 and V6 (data not shown). Fecal calprotectin was quantified in 73 subjects (35 and 38 subjects in Test and Control groups respectively). Calprotectin concentration evolved similarly in both groups with a significant increase from baseline (V2) at the end of *Hp*-treatment (p <0.0001) (Mean (SD): 68.1 (44.8) and 55.4 (37.3) µg/g at V4, in Test and Control respectively), followed by a decrease at V6 (p <0.0001), to reach a similar level between groups, below the baseline values (p <0.05) at V7 (**Figure 5E, F**).

**Figure 5.**
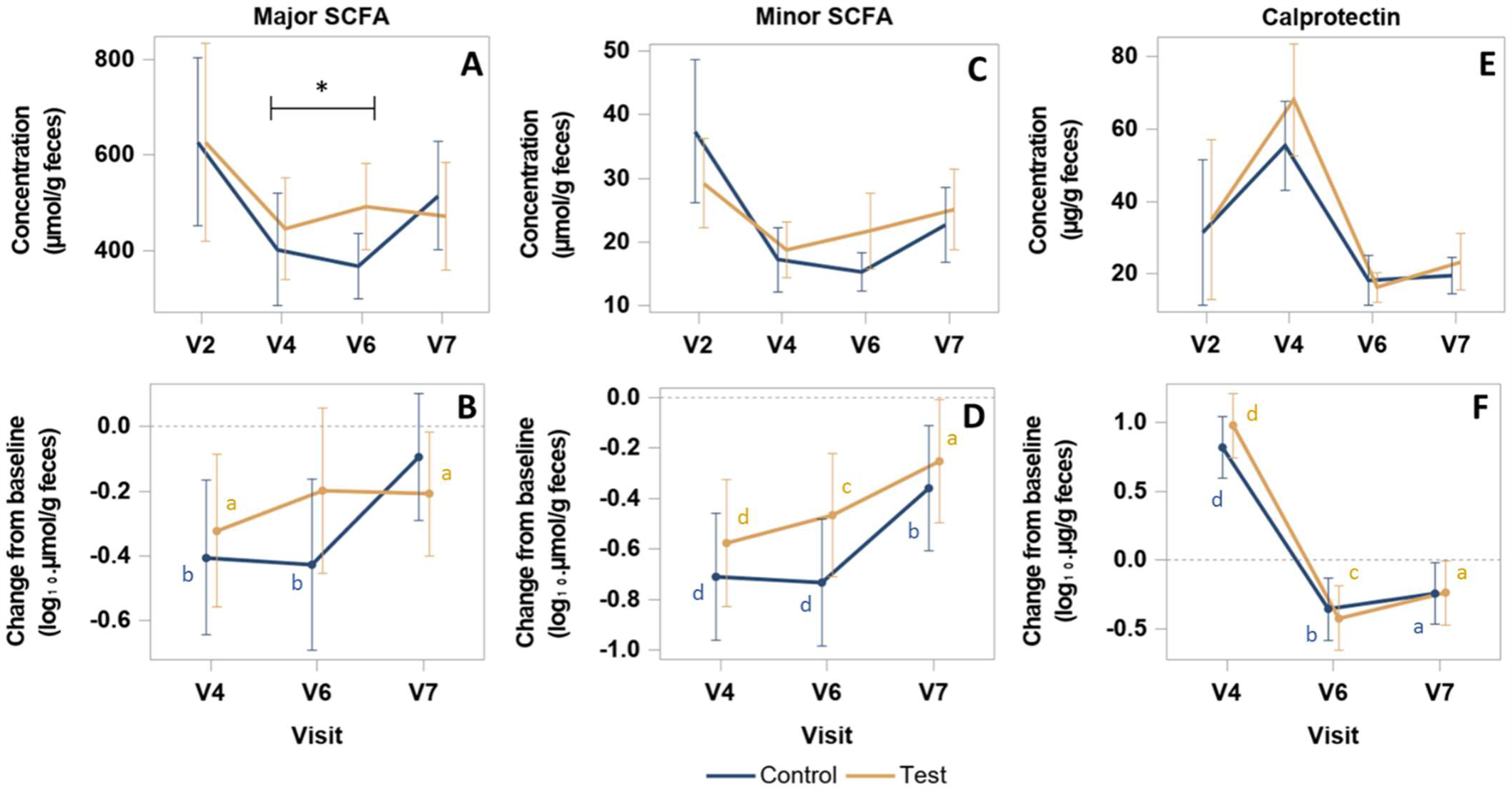
Quantification of fecal major / minor SCFA and Calprotectin. Concentration (A, C, E) and change from baseline (B, D, F) of Major SCFA (acetate, propionate, butyrate) (A, B), Minor SCFA (valerate, caproate, isobutyrate, isovalerate) (C, D) and Calprotectin (E, F). P-values are provided according to Student test: * p <0.05 for comparison between groups of the change of Major SCFA concentration from V4 to V6; ^a^ p <0.05, ^b^ p < 0.01; ^c^ p < 0.001; ^d^ p < 0.0001, for comparison within group of the change from V2 at each visit. SCFA concentrations are expressed in µmol/g of dry feces.

### Safety outcomes

A similar number of AE were reported by 42 (61,8%) subjects in each group (**Additional file 13: Table S4**), mainly headaches, nasopharyngitis, vulvovaginal mycotic infection, dysgeusia and rash. Very few severe AE and one SAE (ankle fracture) were reported in the Control group. AE from which the causality was not established were all qualified as unlikely related to the study products. AE due to *Hp* treatment were reported by 41.2 % of the subjects in each group. For all blood safety parameters, weight or vital signs, no relevant evolution was observed during product intake and follow-up periods and no relevant differences between groups were found at baseline or at any other timepoint.

## Discussion

The present study investigated the effect of a fermented milk product containing *Lp* CNCM I-1518, *Lp* CNCM I-3689 and *Lr* CNCM I-3690 and yogurt strains (Test), on the prevention of AAD and GI symptoms, and on the gut microbiota composition and SCFAs when administered in combination with a standard triple *Hp* eradication therapy.

The Test product did not show a significant effect on AAD occurrence or duration. This can be due to the much lower than expected AAD occurrence observed (1,5% in the Control group instead of the 15% as estimated from former trials) which is most probably due to the young-middle age of a study population with no comorbidities. Other considerations refer to diarrhea not recorded as primary criteria in other studies, or to differences in *Hp* eradication regimens, length of symptom reporting or AAD definitions. The resistance of subjects to *Hp* treatment side effects in our study was also reflected by an absence of GI-symptoms according to GSRS score evolution. In older and more sensitive populations, one of the Test product strains showed an effect in reducing both AAD and CDAD occurrence ^25 26^ with however some exceptions ^33^. The rate of *Hp* eradication was not different between the Test and Control groups in our study and was consistent with previous reports ^34^.

We showed that the 14-day *Hp* eradication treatment induced gut microbiota alteration, reflected by a decrease in total bacterial count and taxonomically-wide changes that persisted up to 28 days following cessation of the treatment, in line with previous studies showing that recovery of gut microbiota following *Hp* triple therapy, might take more time ^12^. Bacteria genera exhibited variable acute and recovery responses, suggesting that a longer follow-up period would have allowed us to study longer-term resilience. Actinobacteria genera were mostly depleted. The durable loss of *Bifidobacterium* is consistent with a previous study in which its abundance was still depleted 3 months following *Hp* triple therapy ^7^. Distinct responses were observed within the *Coriobacteriia* class, with *Slackia, Collinsella* being depleted while *Egghertella*, a known drug-metaboliser ^35^, was enriched.

Intra-subject distance to baseline was lower in subjects consuming the Test product, suggesting a gradual faster rate of recovery of gut microbiota. This contrasts with the study from Suez *et al*, who showed that the fecal microbiota recovery was delayed up to 5 months by the intake of a mixture of 11 lactic acid bacteria and bifidobacteria strains, following broad-spectrum antibiotics intake (ciprofloxacin and metronidazole)^36^. This highlights the variability of gut microbiota response to distinct strains in the context of antibiotics treatment. In the present trial, we observed a bloom of *Escherichia-Shigella and Klebsiella* that persisted in Control but not in Test group up to 28 days after the end of *Hp* treatment, in line with the anti-pathogenic effects of the Test product strains observed in pre-clinical models ^37-39^. Overall, more pronounced differences between product groups were found in *Hp* treated subjects than in healthy subjects ^28^ suggesting that the Test product induces a higher gut microbiota response in a context of altered ecosystem.

The fungal community was previously shown to bloom following antibiotics treatment ^40^. Here, a transient increase in the ratio fungi/bacteria (ITS/16S) following the 14-day *Hp* treatment was observed followed by a reversion towards baseline, which tended to be faster in subjects consuming the Test product. This suggests that the balance of fungi to bacteria was altered probably through additional availability in nutrients and/or direct and indirect effects of some bacteria on the fungal population. Notably, the abundance of *Candida* was transiently increased, in line with a report based on culture method ^41^

In our study, fecal concentrations of all SCFA decreased in both groups at the end of *Hp* treatment and beyond in the Control group until the end of product consumption and then increased after two weeks of follow-up, but without complete recovery of baseline levels. In contrast, SCFA concentration in the Test group showed a significantly faster recovery following cessation of *Hp* treatment, especially for total major SCFA and valerate. Consistently, *Oscillibacter*, for which some species have been shown to produce valerate ^42^, and the butyrate producers *Butyrivibrio* and *Butyricimonas*, were more abundant in the Test group as compared to Control. These results are in line with the capacity of the three strains *Lr* CNCM I-3690, *Lr* CNCM I-3690 and CNCM I-1518 to induce SCFA production and increase the abundance of propionate or butyrate-producers from gut microbiota in different animal or *in vitro* models ^38 39 43 44^. SCFA were shown to create unfavorable conditions for pathobionts ^45^ which may explain the lower abundance of *Escherichia-Shigella* and *Klebsiella* in Test group. The absence of modification of SCFA in blood in the present study suggests that the observed variations of fecal SCFA might reflect a change of SCFA production in the colon, rather than a modification of absorption in blood from the gut.

The transient detection of viable bacteria strains in the feces throughout the Test product consumption period shows the capacity of the three strains to survive in the GI-tract in the presence or absence of *Hp* treatment. Two weeks following cessation of product intake, *Lr* CNCM I-3690 strain was predominantly detected in some subjects. Whether its longer detection is related to higher microbiota permissivity and/or specific characteristics of the strain, such as the expression of pili ^46^, needs further investigation.

A transient increase of calprotectin was also observed in response to *Hp*-treatment in association with gut microbiota alteration. Calprotectin is a marker of gut inflammation that correlates with bacterial infectious diarrhea ^47 48^. Increase of calprotectin production was also associated with PPI intake ^49^ possibly due to bacterial overgrowth as a result of inhibition of acid production in the stomach. In the present study, some genera were durably enriched including AAD-related pathobionts following *Hp*-treatment such as *Escherichia-Shigella* and *Klebsiella*, concomitant to the transient increase of fecal calprotectin. Future metagenomics studies would allow us to determine whether the strains carry virulence genes.

While gut microbiota or SCFA production were significantly altered and calprotectin production transiently increased, only a limited number of subjects reported AAD. A possible explanation could be that gut microbiota alteration was not sufficient to induce a higher occurrence of AAD. In another trial, probiotics reduced microbiota alteration triggered by *Hp* treatment with no improvement of GI symptoms ^50^, which also suggests a possible disconnection between both effects. The low AAD occurrence is also consistent with the calprotectin level, close to normal range in both groups (below 50µg/g feces) as compared to the higher concentrations reported in diarrheic patients ^47 48^ suggesting that *Hp* treatment induced limited or no inflammatory response in our study.

This study has some limitations. First, the low occurrence of AAD did not allow us to observe a product effect on clinical outcomes which limits the conclusion on the clinical relevance of the effects on gut microbiota and SCFA. The trial was also not designed and specifically powered to detect differences in microbiota composition between groups. Finally, subjects without *Hp* treatment were not included which may limit the conclusions as to the cause-effect relationship between medication and the observed modification of study outcomes.

## Conclusion

To our knowledge this is the first study describing the effect of a 14-day *Hp* triple therapy on GI related symptoms, gut microbiota composition (using quantitative microbiome profiling), fungal community, metabolites and host reactive inflammatory markers. *Hp*-treatment was shown to alter all these biological parameters, and the daily consumption of a fermented milk product containing *Lp* CNCM I-3689, *Lr* CNCM I-3690 and *Lp* CNCM I 1518 and yogurt strains, can induce a faster recovery of the microbiota composition and SCFA production and limit the bloom of pathobionts such as *Escherichia-Shigella* and *Klebsiella*. The study product may confer a better resilience following a drug-mediated treatment, therefore protecting the homeostasis of the gut microbiota, and reduce the abundance of potentially pathogenic bacteria.

## Supporting information

CONSORT 2010 Checklist

## Data Availability

16S sequences associated with this project were deposited in EMBL and will be accessible following acceptation of the manuscript for publication in a peer-reviewed journal.

## Abbreviations

AAD: antibiotic associated diarrhea
BSS: Bristol Stool Scale
AE: Adverse Event
Cd: *Clostridioides* difficile
CDAD: *Cd*-associated diarrhea
GI: gastro-intestinal
GSRS: Gastrointestinal Symptom Rating Scale
*Hp*: *Helicobacter pylori*
ICF: Informed Consent Form
*Lp*: *Lacticaseibacillus paracasei*
*Lr*: *Lacticaseibacillus rhamnosus*
PcoA: Principal Coordinate analysis
QMP: Quantitative Microbiome Profiling
SCFA: short chain fatty acids
rRNA: ribosomal RNA

## Declarations

### Ethics approval and consent to participate

The study was approved by the Ethics Committee of Charité – Campus Mitte (Application Number EA1/297/15) of the Charité – Universitätsmedizin Berlin, Germany. All volunteers provided a signed informed consent form.

## Consent for publication

Not applicable.

## Author’s contributions

- Conception and design of the study: E.G, F.S, L.Q, M.D, HS, FB, PM.
- Patient recruitment and study organization: F.S, CK, FW.
- Generation, collection, assembly of data: CK, FW, L.Q, CR, M.P.
- Analysis of and/or interpretation of data: E.G, L.Q, CR, M.D, M.P, CK, FW, HS, FB, PM.
- Drafting or revision of the manuscript: E.G, M.D, L.Q, CR, M.P.
- Critical revision and approval of the final version of the manuscript: F.S, CK, FW, HS, FB, PM.

## Acknowledgements

We would like to thank Aurélie Cotillard for discussions about statistical analysis, Hue Delhomme for support for qPCR-PMA, Marion Ghidi for project management, Nicolas Bouchet for overall analytical support.

## Supplementary material

**Additional file 1: Methods**. Additional methods

**Additional file 2: Figure S1**. Decision tree for diagnosis of *Hp* infection and subjects screening

**Additional file 3: Table S1**. Bacterial strain counts in Test product during the shelf life

**Additional file 4: Figure S2**. Subject flowchart

**Additional file 5: Results**. Additionnal results

**Additional file 6: Table S2**. Number of days of GI symptoms

**Additional file 7: Figure S3**. GSRS

**Additional file 8: Figure S4**. Quantification of Test product strains in feces

**Additional file 9: Figure S5**. UniFrac beta-diversity

**Additional file 10: Figure S6**. Global gut microbiota response to *Hp* treatment by Quantitative Microbiome Profiling

**Additional file 11: Table S3**. Differential abundance of all genera across the study based on QMP, expressed as Fold change and adjusted p values

**Additional file 12: Figure S8**. Quantification of fecal individual SCFA

**Additional file 13: Table S4:** Adverse events

**Additional file 14: Figure S7**. Principal Coordonate analysis of Bray-Curtis on ITS

## Additional file 1: Methods

**Subject Eligibility Criteria**

## Inclusion criteria

- 1: Subjects who read and signed the Study Informed Consent Form.
- 2: Adult male/female [18, 65] years old.
- 3: Subjects with a body mass index (BMI) as follows: 19 ≤ BMI ≤ 30 kg/m^2^
- 4: Subjects positive for *Helicobacter pylori* (*Hp*) infection and symptomatic due to *Hp* infection.

Diagnosis of *Hp* infection based on positive ^13^C-Urea Breath test and at least one positive test among Urease test and Histological test for *Hp*-gastritis

*Subjects symptomatic due to H. pylori infection were intended here as subjects with at least one or more of the following dyspeptic symptoms:*

a. *Bothersome postprandial fullness: Uncomfortably full after regular sized meal, more than 1 day/week*.
b. *Early satiation: Unable to finish regular sized meal, more than 1 day/week*.
c. *Epigastric pain: Pain or burning in middle of abdomen, at least 1 day/week*.

*Histology of gastric biopsies were performed in positive 13C-Urea Breath test patients with dyspeptic symptoms*

-5: Subjects with an indication, as stated by a Gastroenterologist, for the eradication of *Helicobacter pylori* with a *Helicobacter pylori* eradication therapy as follows: Proton Pump Inhibitor (pantoprazole 40 mg twice daily), clarithromycin 500 mg-twice daily, amoxicillin 1000 mg twice daily, for 14 days.
-6: For female: If of child bearing potential, female subjects must be using or complying with one of the following medically approved methods of contraception such as, but not exclusively:
  a. Oral birth control pills (at least 1 full monthly cycle prior to study product administration);
  b. Intra-uterine device (IUD);
  c. Double barrier methods (such as condoms and spermicide);
  OR Female subject must be postmenopausal for at least 12 months prior to trial entry or surgically sterile (i.e. hysterectomy, bilateral oophorectomy or bilateral tubal ligation).
-7: Subjects who could and were willing store a maximum of 32 study product bottles (100g each) at any one time in his/her fridge (between +2°C and +8°C).
-8: Subjects who accepted an alimentary restriction of products including yoghurts, fermented dairy products with probiotics, over-the-counter medication containing probiotics, starting at Day 0 until Last study visit (at least 42 days).
-9: Subjects who appreciated dairy products and multi-fruit flavour and who consumed 2 bottles per day of study product for 28 consecutive days.
-10: Subjects who were willing to provide stool and blood samples throughout the duration of the study.
-11: Subjects who were willing to complete ePRO and had access to internet.

## Exclusion criteria

### General

-1: Female subjects with a positive pregnancy test (based on serum test), or planning to become pregnant during the study or breast-feeding women.
-2: Subjects enrolled in another interventional clinical study in the last 4 weeks or in an exclusion period following participation in another clinical trial.
-3: Subjects who reported a foreign travel in the previous 7 days or who planned a foreign travel during the study in a country at risk for contracting diarrhea, including Asia (except for Japan), Middle East, Africa, Mexico, and Central and South America.
-4: Subjects not able to answer questionnaires by writing whatever the reason.
-5: Subjects with a loss of personal liberty, by administrative or judicial decision.
-6: Subjects who were major but with a legal guardian.
-7: Subjects in a situation which in the investigator’s opinion could interfere with optimal participation in the present study or could constitute a special risk for the subject.
-8: Subjects who had a history of alcohol abuse in the 6 months preceding the study.

### Symptoms

-9: Subjects having diarrhea within the preceding 4-weeks (defined as any episode of stool of 5-7 types in Bristol Stool Scale).

### Medical history

#### Global

-10: Subjects with severe life-threatening illness.
-11: Subjects presenting a severe evolutive or chronic pathology (e.g. cancer, tuberculosis, Crohn’s disease, cirrhosis, multiple sclerosis, Type I diabetes…).

#### Immune disorders

-12: Immune-suppressed subjects (for congenital reasons or acquired due to HIV infection, malignancy or its treatment, steroids – endogenous excess or exogenous, post-transplantation or patients receiving cyclosporin, or any clinical condition compromising the patient’s immune function).

#### Gastro-intestinal pathologies

-13: Subjects with at least one alarm feature including bleeding, anemia, unexplained weight loss, dysphagia, odynophagia, recurrent vomiting, previous malignancies in the gastrointestinal tract.
-14: Subjects with benign peptic ulcer or pre-malignant or malignant lesions based on gastroscopy exams within the context of *H. pylori* detection.

#### Note

-Duodenal ulcer was not a non-inclusion criteria.
-For all subjects presenting at least one dyspeptic symptom (i.e, bothersome postprandial fullness or early satiation or epigastric pain), a gastroscopy is proposed systematically.
-15: Subjects presenting an infection of the of the gastrointestinal tract.
-16: Subjects presenting a severe evolving or active pathology of the gastrointestinal tract such as cancer, inflammatory bowel syndrome, inflammatory bowel disease, Crohn’s disease or ulcerative colitis, diverticular disease, biliary disease, chronic liver disease (based on blood transaminase analysis), any clinical condition affecting the pancreas including acute and chronic pancreatitis, history of metabolic disease (malabsorption, celiac disease) with the exception of dyspepsia and infection related to *Helicobacter pylori*.
-17: Subjects with any past severe gastro-intestinal or metabolic pathology with the exception of appendicitis.
-18: Subjects with a history of *Hp* eradication therapy.

#### Cardiac, renal and surgery issues

-19: Subjects with cardiac insufficiency.
-20: Subjects with a history of angina pectoris or other clinically significant cardiac disease.
-21: Subjects with artificial heart valves (cardiac valvular prosthesis).
-22: Subjects with a history of endocarditis.
-23: Subjects with a history of rheumatic fever.
-24: Subjects with a congenital heart defect.
-25: Subjects with non-controlled hypertension.
-26: Subjects having had any surgery or intervention requiring general anaesthesia in the last 4 weeks, or who had planned surgery or an intervention requiring general anaesthesia in the course of the study.
-27: Subjects who had any dental surgery in the last 4 weeks, or that had planned dental surgery in the course of the study.
-28: Subjects who had gastro-intestinal surgery in the last 3 months or planned surgery in the course of the study.
-29: Subjects with renal insufficiencies based on blood creatinine analysis.

#### Treatments

-30: Subjects with allergy or hypersensitivity against the medication for the *Hp* eradication therapy.
-31: Subjects with antibiotic or intestinal antiseptic treatment during the previous 2 months.
-32: Subjects with H2-receptor antagonists or PPI treatment in the last 2 weeks.
-In case of use of the above treatments in the past two weeks, the subjects were asked to stop the treatment and come back 2 weeks later for *Hp* infection diagnosis and inclusion in the study (unless ineligible for other criteria).
-33: Chronic use of laxatives or anti-diarrheal.
-In case of use of the above treatments in the past two weeks, the subjects were asked to stop the treatment and come back 2 weeks later for inclusion in the study.
-34: Subjects currently receiving immunosuppressants or chemotherapy.
-35: Subjects taking any treatment aimed at weight management or any form of treatment likely to interfere with metabolism or dietary habits.

#### Dietary issues

-36: Subjects with any diagnosed food allergy.
-37: Subjects with allergy or hypersensitivity to any component of the study products (e.g.: allergy or hypersensitivity to milk proteins or lactose).
-38: Subjects with eating disorders (anorexia, boulimia).
-39: Subjects with special medicated diet (due to obesity, anorexia, metabolic pathology).
-40: Subjects under artificial nutrition in the last 2 months.

#### Breath Test

^13^C-Urea Breath Test was carried out using HeliFANplus device (Fisher Analysen Instrumente GmbH) ^1^, before and 30 minutes after ingestion of an oral substrate dose of 75mg of [^13^C] urea (99%), dissolved in 250mL of an acidic drink.

#### Fecal sample collection

A total of 558 fecal samples was collected in the study from 135 (67 in Test and 68 in Control group) subjects at 4 time points (Figure 1). Fresh fecal samples were collected and kept up to 24h at 2-8°C in anaerobic conditions (Genbag Anaerobic, Biomerieux), before being aliquoted and stored at -80°C. All samples were then processed for total bacterial count (flow cytometry) and for DNA extraction in view of microbiota profiling. For SCFA, calprotectin, and Test product strains viability analyses, samples from a subgroup of subjects were analyzed.

#### Clostridioides difficile detection in feces

Test for *Clostridioides difficile* (Cd) in feces followed a two-step strategy as recommended by the European clinical guidelines ^2^. Detection was based on Glutamate dehydrogenase (GDH) and *Cd*-Toxin A and B tests (*Cd*. Quick Check Complete ®, Abbott, US). In case of GDH-/Toxin+ or GDH+/Toxin-results, Polymerase Chain Reaction (PCR) test was used to confirm the presence or absence of a toxigenic strain. *Cd* detection tests were performed in stools collected at day 0, day 28 and in case of AAD (in two samples, one collected on the first day of AAD and one after the end of AAD, i.e. after two consecutive days with stools with BBS type <5).

#### Quantification of total and viable Test product strains in feces

Strain quantification was performed in 48 subjects, including 40 subjects randomly selected from the FAS population and the 8 subjects who reported an AAD-definition 2. Results are expressed as Total cell count (both live and dead cells), Viable cell count, Viability loss (Total cell count – Viable cell count) and Viability rate ([Viable cell count/Total cell count]×100) at each planned visit. Total count of three strains from Test product was assessed using a real time quantitative PCR (qPCR) while the count of viable strains was determined through a qPCR combined with propidium monoazide labelling (PMA, Sigma Aldrich, France), that can differentiate live from dead bacteria. Each fecal sample was analyzed in three conditions: (1) qPCR without PMA treatment for total bacteria, (2) PMA-qPCR for viable bacteria and (3) qPCR heat treatment at 95°C/20 min and PMA treatment for PMA efficacy control. PMA was added to 2mg of feces at a final concentration of 100 µM, incubated 5 min in dark at room temperature (RT) with occasional mixing. All samples were light exposed for 15 min, mixed every 3 min at RT using a PhAST Blue Photoactivation System (Geniul, Spain). DNA extraction was performed as previously described ^3^. PCR reactions contained an internal control for presence of any PCR inhibition (Eurogentec, Belgium). qPCR run was performed on a 384-well plate (Eppendorf®) with triplicate wells for each sample. qPCR reactions included 2×QuantiFast Multiplex PCR Mastermix (QIAGEN, France), 1 μL extracted fecal DNA, 500 nM for each primer ^3^ and 200 nM for each Taqman probe. qPCR amplification was programmed with an initial denaturation at 95 °C for 5 min, followed by 40 cycles of 95 °C/30 s and 60 °C/30 s.

### Gut Microbiota

#### Flow cytometry

Sample preparation and flow cytometric analysis of the samples was done according to Vandeputte et al. with some minor modifications ^4^. For cell counting, 0.1g of fecal sample was weighed inside a sterile falcon tube using an analytical balance. To this material, 10 mL of sterile PBS was added, and the samples were vortexed thoroughly. Afterwards, the samples were stored for at least 2 hours in the fridge (4°C) to allow proper homogenization of the samples. Subsequently the samples were again vortexed thoroughly. Upon complete homogenization the samples were diluted 100 times in sterile PBS and filtered through a sterile filter (pore size 5 μm). Finally, 50 µL of these 100 times diluted samples was added to 445 µL sterile PBS. The microbes present in these final dilutions were stained with SYTO 24 green fluorescent nucleic acid stain by adding 5 µL of 0.1 mM-stock solution of SYTO 24 to the samples. Finally, the samples were incubated in the dark for 15 min at 37°C prior to flow cytometric analysis. Stained samples were analyzed on a BD FACSVerse. Proper PMT voltage, threshold, and gating settings separated the stained microbes from sample debris and signal noise. The samples were run at medium flow rate (60 µL/min) and events were recorded in a SSC-H:FITC-H dot plot. A threshold of 200 was applied on the FITC-H channel. After flow cytometric analysis, data were analyzed and processed in Flow jo V10.

#### DNA extraction, 16S and ITS2 sequencing

DNA was extracted from 200 mg frozen stool via chemical and mechanical lysis (Fastprep® FP120 [ThermoSavant]) as described previously ^5^. 50 ng of DNA was amplified following the 16S Metagenomic Sequencing Library Illumina 15044223 B protocol (ILLUMINA) using V3-V4 primers for 16S rRNA gene ^6^and ITS2 ^7^. The samples were loaded into flow cells in an Illumina MiSeq. 300PE Sequencing Platform in accordance with the manufacturer’s instructions. 539 fecal samples were kept for downstream 16S rRNA analyses (135 subjects, (67 in Test and 68 in Control group) for Control) and 374 samples for ITS analyses (97 subjects, N=49 for Test and N=48 for Control).

#### Bioinformatic analysis

Raw sequences, forward and reverse, were merged in order to obtain the complete sequence using the ‘pear v0.9.6’ software. The amplification primers from the sequences obtained in the sequencing step were trimmed to reduce the bias in the annotation step, with ‘cutadapt v 1.8.1’ and parameters by default. Once the primers removed, sequences smaller than 200 nucleotids were removed from the analysis. After obtaining the clean complete sequences, a quality filter was applied to them to delete poor quality sequences. Those bases in extreme positions that did not reach Q20 (99% well incorporated base in the sequencing step) or a greater quality score were removed. Subsequently, sequences whose average quality did not surpass the Q20 threshold, as a mean quality of the whole sequence, were also deleted. The resulting sequences were inspected for PCR chimera constructs that may occur during the different experimental processes. Chimeras were removed from further analysis. Analyses were performed using QIIME (v. 19). After filtering for quality, a mean of 99,437± 36,973 sequences per sample were retained. Reads were clustered into operational taxonomic units (OTUs; 97% identity threshold) using VSEARCH, and representative sequences for each OTU were aligned and taxonomically assigned using the SILVA database (v. 119) and the Unite database (v. 7.2 (ITS). To characterize diversity, rarefaction was used to obtain 22,000 sequences per sample. Alpha diversity (within samples) was assessed using Shannon index) and Simpson’s Reciprocal index for 16S rRNA and using Shannon index for ITS). The ITS/16s ratio was computed for Shannon index ^8^. Beta diversity was represented using Bray-Curtis dissimilarity and UniFrac distances (weighted and unweighted) for 16S rRNA and Bray-Curtis dissimilarity for ITS.

#### SCFA and Calprotectin dosage

SCFA and Calprotectin were analyzed in subgroups of the last 61 and 73 subjects randomized respectively. Calprotectin concentration in feces was measured by ELISA (PhiCal Calprotectin ELISA, Immungiagnostik AG, Germany). SCFA concentration in stools (per gram of dry feces) and in serum was measured by headspace gas chromatography mass spectrometry (GC-MS) (column Rx-624Sil MS, RESTEK, France).

#### Safety parameters

Safety assessment was performed by following vital signs (systolic and diastolic blood pressure, heart rate, body temperature) and anthropometry (body height, weight and Body Mass Index) at days -45 (up to), 0, 7, 14, 21, 28 and 42, blood analyses of partial hemogram (red blood cells, leucocytes, haemoglobin, haematocrit, neutrophils, eosinophils, basophils, lymphocytes, monocytes and platelets), glucose, insulin, Insulin Resistance (HOMA-IR), cholesterol (total, HDL, LDL) and triglycerides at days 0, 28 and 42, hepatic enzymes (ALT, AST, γGT) and creatinine at days -45 (up to), 0, 28 and 42. The occurrence and the number of spontaneously reported adverse events were assessed throughout the study as well as their relationship to the study product or the *Hp-*treatment, their intensity, seriousness, and the appropriate action taken (interruption of product intake or *Hp*-treatment or subject withdrawal).

#### Procedure

Subjects underwent a physical examination and vital signs were recorded at each visit. At V2, subjects reported physical activity habits on a usual week by answering the Short last 7 days self-administered version of the Physical Activity Questionnaire (IPAQ) (revised august 2002 version) ^9^ and smoking habits on the last month (expressed in number of smoked cigarettes per week, using the following equivalences: 1 pipe = 5 cigarettes; 1 small cigar or cigarillos = 2 cigarettes; 1 big cigar = 5 cigarettes). Subjects dietary habits during the past month was reported at V2 and V6 at investigation site through a Food Frequency Questionnaire (FFQ) ^10^, adapted for diet reporting of the past month. FFQ outputs were converted in daily amounts of nutrient intake using software developed by the German Institute of Human Nutrition (DIFE), Potsdam. Subject alcohol consumption was reported weekly at all visits and converted to the average number of alcohol units per week (one unit= 10 grams of pure alcohol). In a personal e-diary, through electronic Patient Reported Outcomes (ePRO) software (Patient Cloud, Medidata), subjects daily reported their compliance with the *Hp* eradication therapy, with the study product consumption and with the dietary restrictions, in the corresponding periods, as well as any adverse event, concomitant medication and nutritional supplement intake from V2 to V7. Between V2 and V6, subjects also used ePRO for daily reporting of all passages of stool and their consistency (from 1 to 7 as based on BSS ^11^ and of main GI symptoms, and for filling GSRS questionnaire ^12^ once a week. E-diary was remotely monitored for completion by the investigator or authorized staff on a daily basis and weekly during visits. ^13^C-urea breath test was performed at V1 for *Hp* infection diagnosis process and at V7 (not earlier than 28 days after the last *Hp* treatment dose) to check *Hp* eradication. The study was performed in accordance with the protocol with no change to trial outcomes during the course of the trial. Source data verification was performed by monitors appointed by a third-party contractor (Pharmaceutical Product Development, llc).

### Data Monitoring Committee and Interim analysis

#### DMC organisation

DMC was composed of five independent experts in gastro-enterology and treatment of *Hp* infection, in pediatrics, gastro-enterology, and probiotics, in public health, infectious diseases and management of nosocomial infections, in development of PRO instruments and in Statistics. Details of DMC organization, responsibilities, data flow, decision algorithm and communication were provided in a DMC charter that was signed together with final Statistical Analysis Plan before the interim database lock. The study staff (except the product supply supervisor) including the sponsor statistician remained blinded until the end of the study.

#### Stopping rules at interim analysis

Different scenarios and decision rules were presented to the DMC members to guide the recommendation to stop or to continue the study after the interim analysis. For each scenario, recommendations were based on a sample size re-estimation through the adaptative approach of the Conditional Power (CP) proposed by Mehta and Pocock. For each AAD definition, CP were computed based on data observed on the first 104 evaluable subjects for primary efficacy outcome (Definitions 1 and 2 of AAD occurrence). The following decisions algorithm was carried out for each of the Definition (X = 1 or 2) defined:

-If CP < 30% then the primary outcome for Definition X of AAD was futile
-If 30% ≤ CP ≤ 60% then the sample size required to increase the CP to 80% was computed and the study could continue as a POC for Definition X with a sample size minimum of 550 evaluable subjects
-If CP > 60% then the study continued as a Proof Of Efficacy (POE) for Definition X of AAD with a sample size of 550 evaluable subjects to reach 80% power.

#### Interim analysis

The interim analysis was performed when 104 subjects could be evaluated, and was reviewed by the DMC. As only one subject experienced AAD episode on AAD definition 1, the decision algorithm was based on AAD definition 2. Finally, Definition 2 of AAD occurrence was too low and primary outcome judged as futile. No sample size re-estimation was made. The study sponsor followed the DMC recommendation to discontinue the trial after interim analysis due to futility of clinical efficacy endpoints. As recruitment continued during the interim analysis, 136 subjects were randomized in total for which all analyses had been completed and are presented here.

### Statistical analysis

#### SCFA, Calprotectin, quantification of Test product strains

The *Hp* treatment effect on change of biological parameters was assessed with a repeated measures linear mixed model. For Calprotectin and each SCFA, three covariance structures (unstructured, autoregressive and compound symmetry) were investigated and the best model with the lowest Akaike Information Criteria was retained. Normality of residuals was verified with skewness and kurtosis, for both statistics a [-2; 2] interval was considered as acceptable to state as the normality. If the assumption of normality was not met, a log-transformation was applied, and the same repeated linear model was used. Due to exploratory context, no alpha adjustment for multiple testing strategies was planned. For the quantification of Test product strains in feces, for each strain quantified, the effect of time points (V4, V6 and V7) on total and viable cell count was assessed using a non-parametric Friedman test. A 2 by 2 comparison (Wilcoxon signed-rank test) for time points V4 vs. V6 and V6 vs. V7 was performed with a Bonferroni correction for multiple testing. Due to a higher rate of missing data at V7 observed for viability loss and viability percentage, the effect of time points (V4 and V6) was assessed with a Wilcoxon test.

#### Gut microbiota and mycobiota

For beta diversity, associated tests of multivariate homogeneity of group dispersion were computed as heterogeneity can bias significance in PERMANOVA. Principal Coordinate Analyses were performed to complement PERMANOVA. An intra-subject analysis was performed comparing V4, V6, and V7 with the corresponding baseline (V2) using Mann-Whitney U test. DESeq2 model was set to identify differentially abundant genera between Group and Visit while considering the microbiota composition at baseline (V2) and controlling for differences within individuals, as detailed in the DESeq2 tutorial. For QMP, the normalization step was thus omitted, and the model built on the QMP-normalized values divided by 100 for R to deal with large counts. P-values were adjusted for each parameter in each dataset and alpha risk was set to 0.05.

## Additional file 5: Results

### Clinical outcomes

Results on alternative AAD definition, time to first event and AAD duration (Definition 1 and 2) did not show any significant difference between groups either (data not shown). A slightly higher mean number of days with GI symptoms was observed in the Test as compared to the Control group (mean (SD) of 9.5 (8.0) vs. 8.4 (8.5) days), mainly due to bloating, but the difference was low in proportion and no conclusion can be driven on any product effect (**Additional file 6: Table S2**). No relevant difference between groups was observed either for the Total GSRS score or the GSRS scores by dimension at each kinetic point (**Additional file 7: Figure S3**) or as a change from baseline at each visit. Total and dimensions scores globally decreased, albeit slightly (<1-point score evolution), all along the study between V2 and V6, with the same trends in both groups. Diarrhea dimension score was the only one showing a slight increase between baseline (V2) and V3 or V4, similarly in both product groups.

### Test product strains viability in feces

The viability of the three strains *Lp* CNCM I-3689, *Lr* CNCM I-3690 and *Lp* CNCM I-1518 was assessed in the feces of 48 subjects (25 in Test, 23 in Control group) (**Additional file 8: Figure S4**). Some samples from Control group at V4 or V6 and from Test group at V2, showed a detectable signal in Total cell count of strains, mainly *Lp* CNCM I-1518, in less than 5% of the tested samples overall, in the absence of Test product intake, representing therefore a low proportion of possible false positive detection. At V4, after 2 weeks of Test product intake and concomitant *Hp*-treatment, total cell (live and dead) counts reached 8.1 to 8.3 log_10_ cells/g feces depending on the strain. Viable cell count ranged from 6.5 to 7.1 log_10_ /g feces, with the highest count found for the *Lp* I-1518 strain (**Additional file 8: Figure S4**). The associated viability rate varied accordingly from 2.2% for *Lr* I-3690 to 2.3% for *Lp* I-3689 and 9.4% for *Lp* I-1518, corresponding to a viability loss of 1.7, 1.6 and 1 log_10_ respectively. At V6, for the three strains, at the end of Test product consumption period, no significant change was observed for either the total or viable cell count as compared to V4 when *Hp* treatment ceased (p>0.05). The viability rate and resulting viability loss were also not significantly different between V4 and V6 for *Lr* I-3690 and *Lp* I-3689 whereas a statistically significant decrease of the viability rate (from 9.4 to 3.5%, Friedman Rank Test p<0.05) and increase of viability loss (from 1 to 1.5 log10, Friedman Rank Test p<0.01) was observed for *Lp* I-1518. At V7, two weeks after the end of the last Test product intake, both total and viable bacteria count significantly decreased for all three strains as compared to V6 (Wilcoxon signed-rank, p<0.001) to an undetectable level (LOQ), except for *Lr* I-3690 in 5 and 2 subjects (20% and 8% of subjects) and for *Lp* I-3689 strain in 1 and 1 subject (4% and 4% of subjects) respectively.

### Gut mycobiota response to Hp treatment

At baseline, Ascomycota was the major phylum (median of 60% of total mycobiota). Most dominant assigned fungal genera included *Saccharomyces, Penicillum, Candida, Aspergillus and Torulaspora*. Beta-diversity measured by Bray-Curtis dissimilarity showed that there was a global shift in mycobiota composition at V4 compared to baseline (PERMANOVA adjusted p = 0.005) and a gradual but incomplete recovery (PERMANOVA, adjusted p = 0.015) **(Additional file 14: Figure S7)**. Test and Control were not different at any timepoint. Intra-subject diversity as distance to baseline was not different between group at any timepoint or along time.

**Additional file 11: Table S3.** Differential abundance of all genera across the study based on QMP, expressed as Fold change and adjusted p values-**provided**

**Additional file 2:**
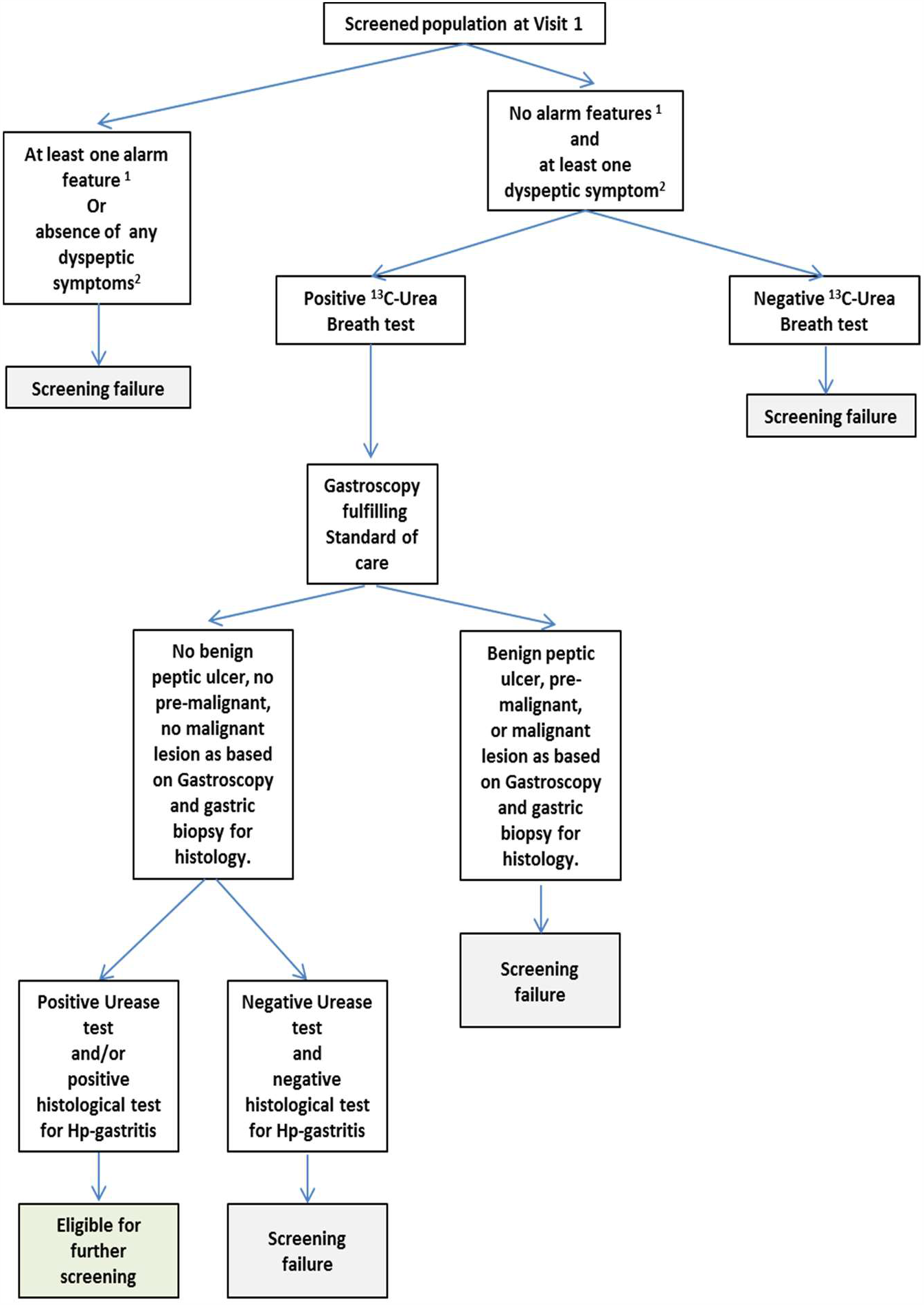
Figure S1. Decision tree for diagnosis of *Hp* infection and subject screening. ^1^ : Alarm features include: bleeding, anemia, unexplained weight loss, dysphagia, odynophagia, recurrent vomiting, malignancies in the gastrointestinal tract ^2^ : Dyspeptic symptoms : bothersome postprandial fullness, early satiation, epigastric pain.

**Additional file 4:**
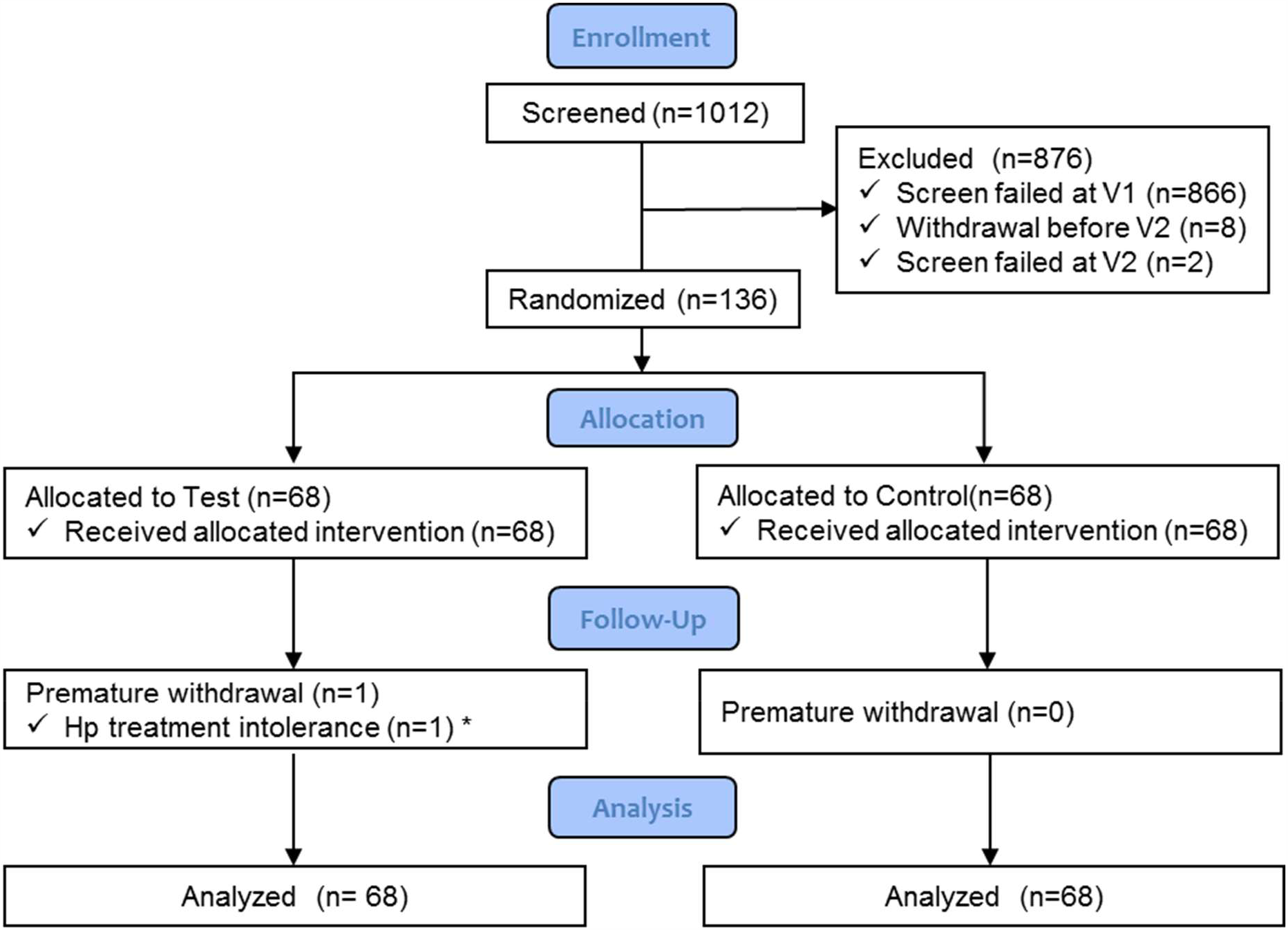
Figure S2. Subject flowchart. *subject was analyzed in the FAS population

**Additional file 7:**
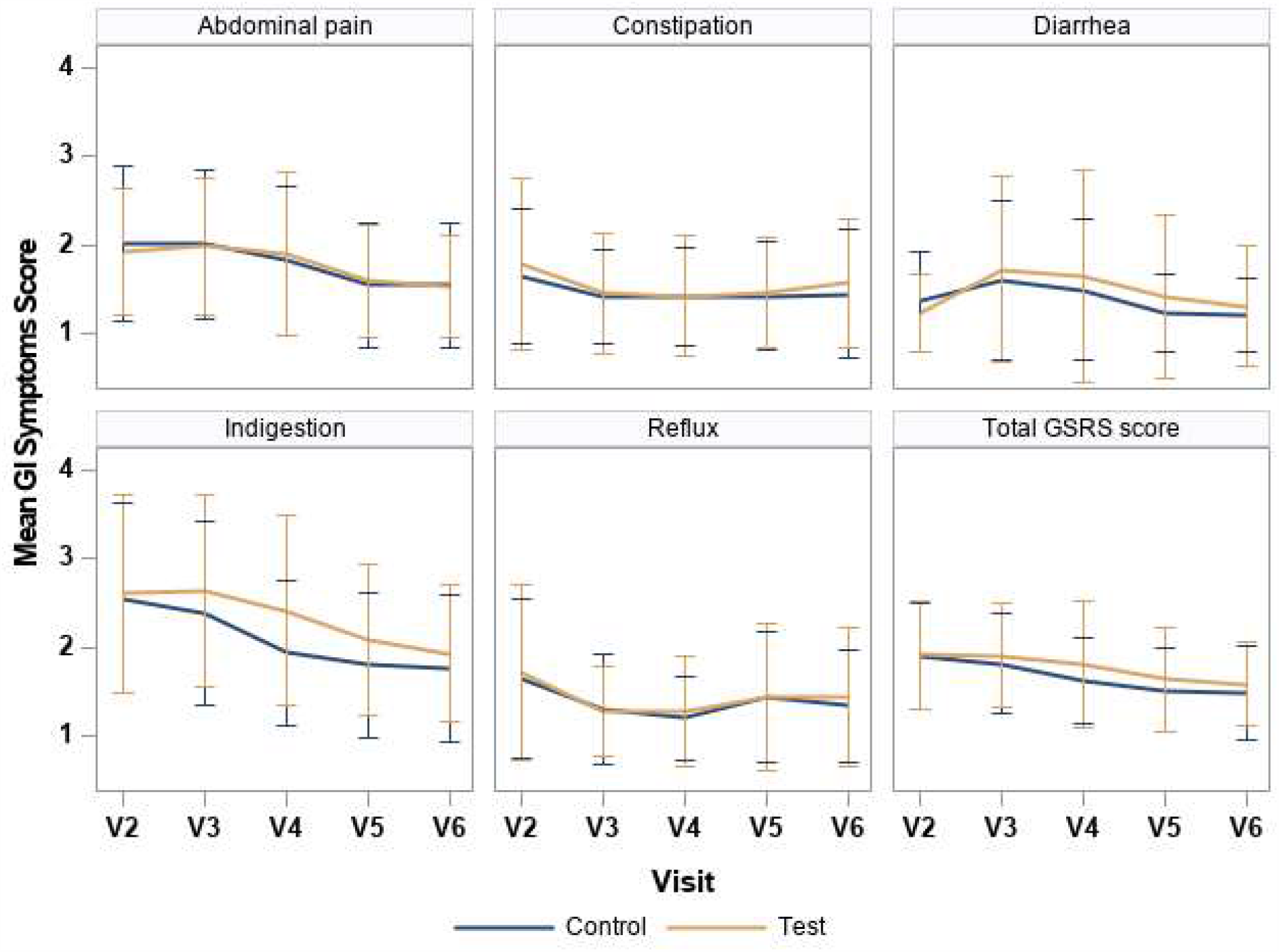
Figure S3. GSRS. Total GSRS score and scores by each GSRS dimension per visit.

**Additional file 8:**
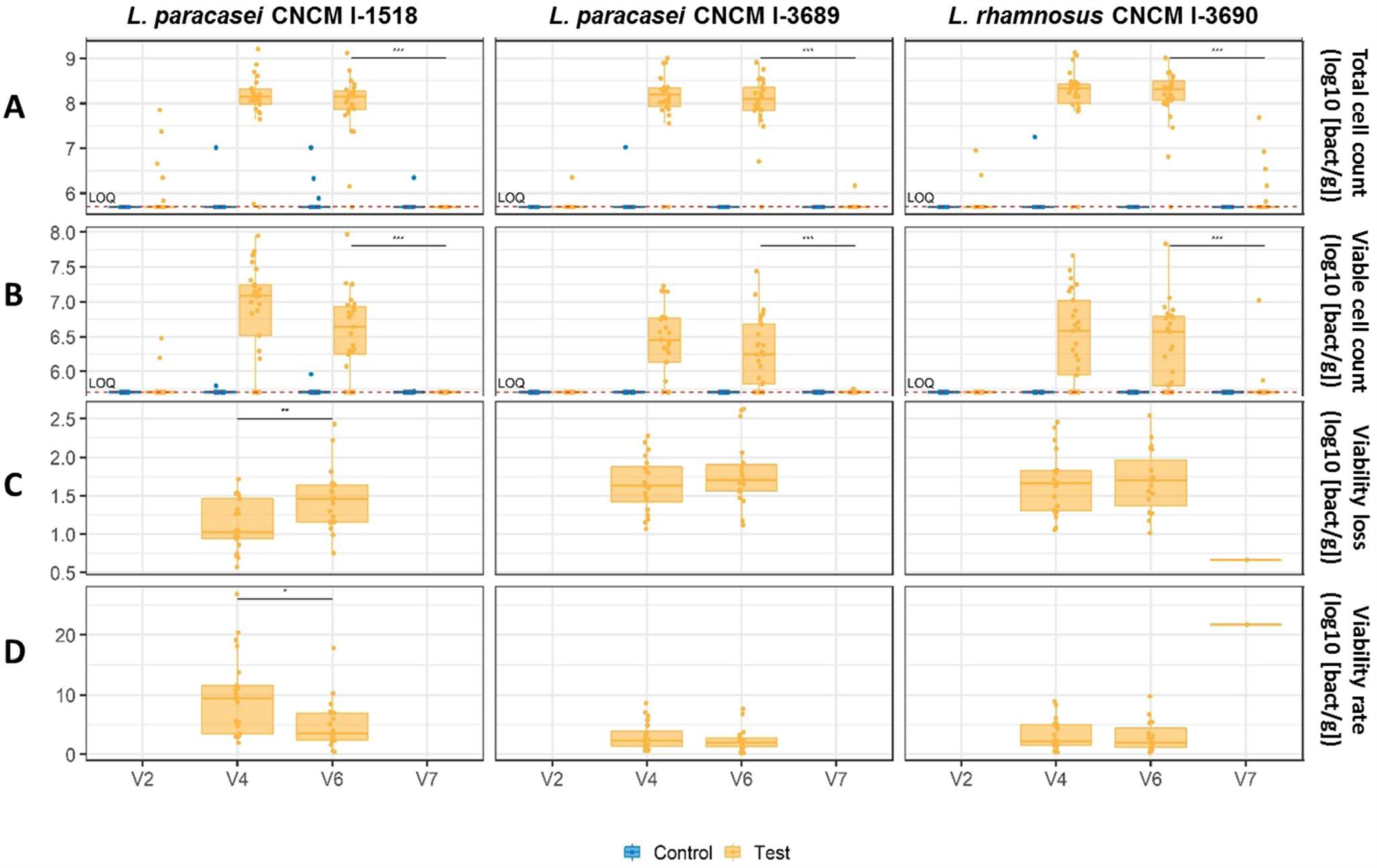
Figure S4. Quantification of Test product strains in feces. (A) Total (live and dead) cell count,(B) Viable cell count, (C) Viability loss (Total cell count – Viable cell count) and (D) Viability rate ([Viable cell count/Total cell count]×100), per visit. For Total and Viable cell count, data below the limit of quantification (LOQ) were imputed at the LOQ (5.7 log_10_ [bacteria cell number/g feces]). Imputed values are used for V4, V6, V7 data and non-imputed values for analyses of changes between visits (V6-V4, V7-V6). For Viability loss and Viability rate, when Total or Viable cell counts were below LOQ, data were not imputed and considered as missing. P-values are given for comparison between visits in Test group and after Bonferroni correction for Total and Viable cell count variables (*= <0.05, **=<0.01; ****=<0.0001). Total cell count, Viable cell count and Viability loss are expressed in log_10_ [bacteria cell number/g feces])(log_10_ [bact/g]).

**Additional file 9:**
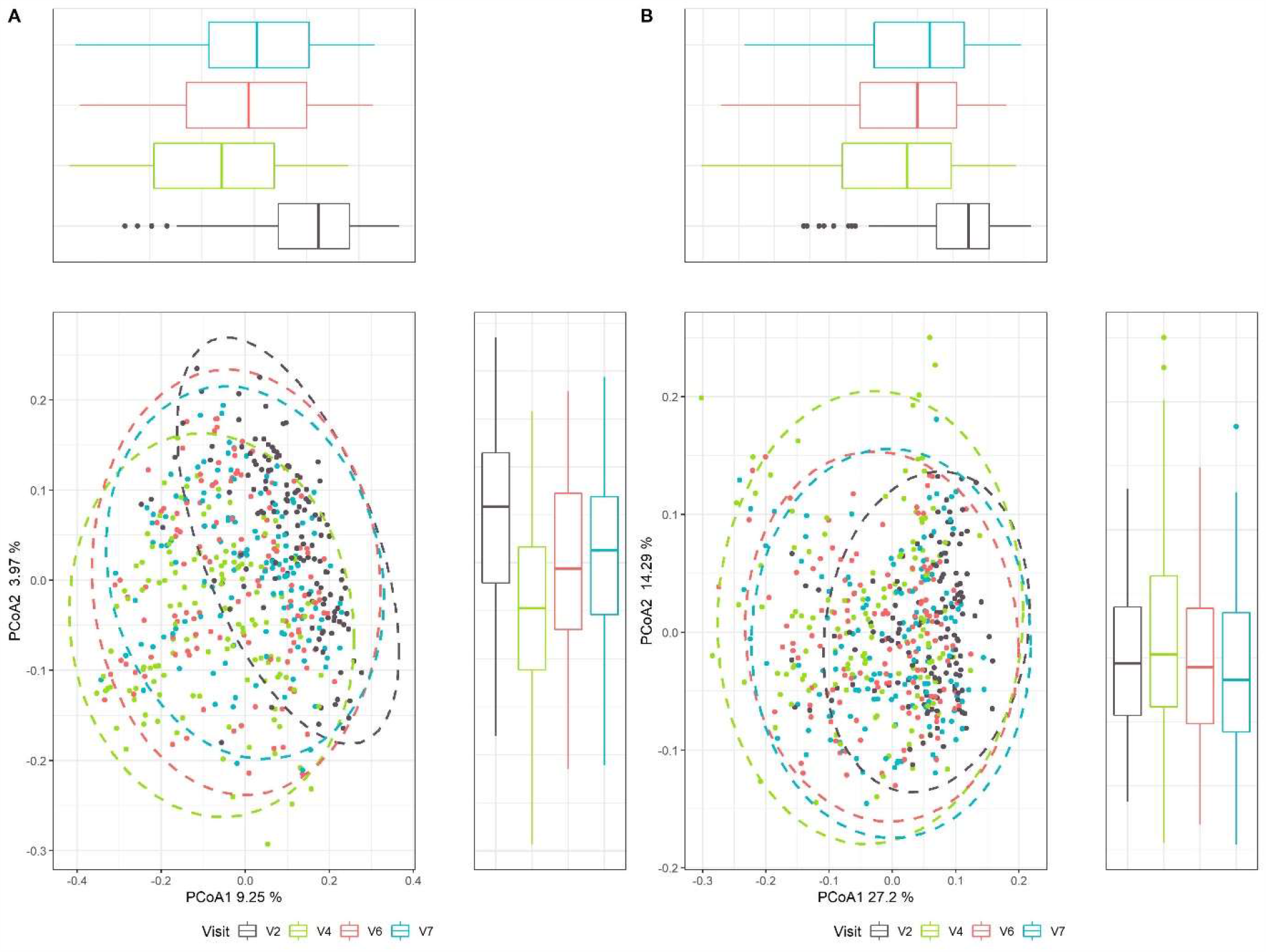
Figure S5. UniFrac beta-diversity. (A) Principal Coordinate analysis (PcoA) of unweighed UniFrac (B). weighted UniFrac.

**Additional file 10:**
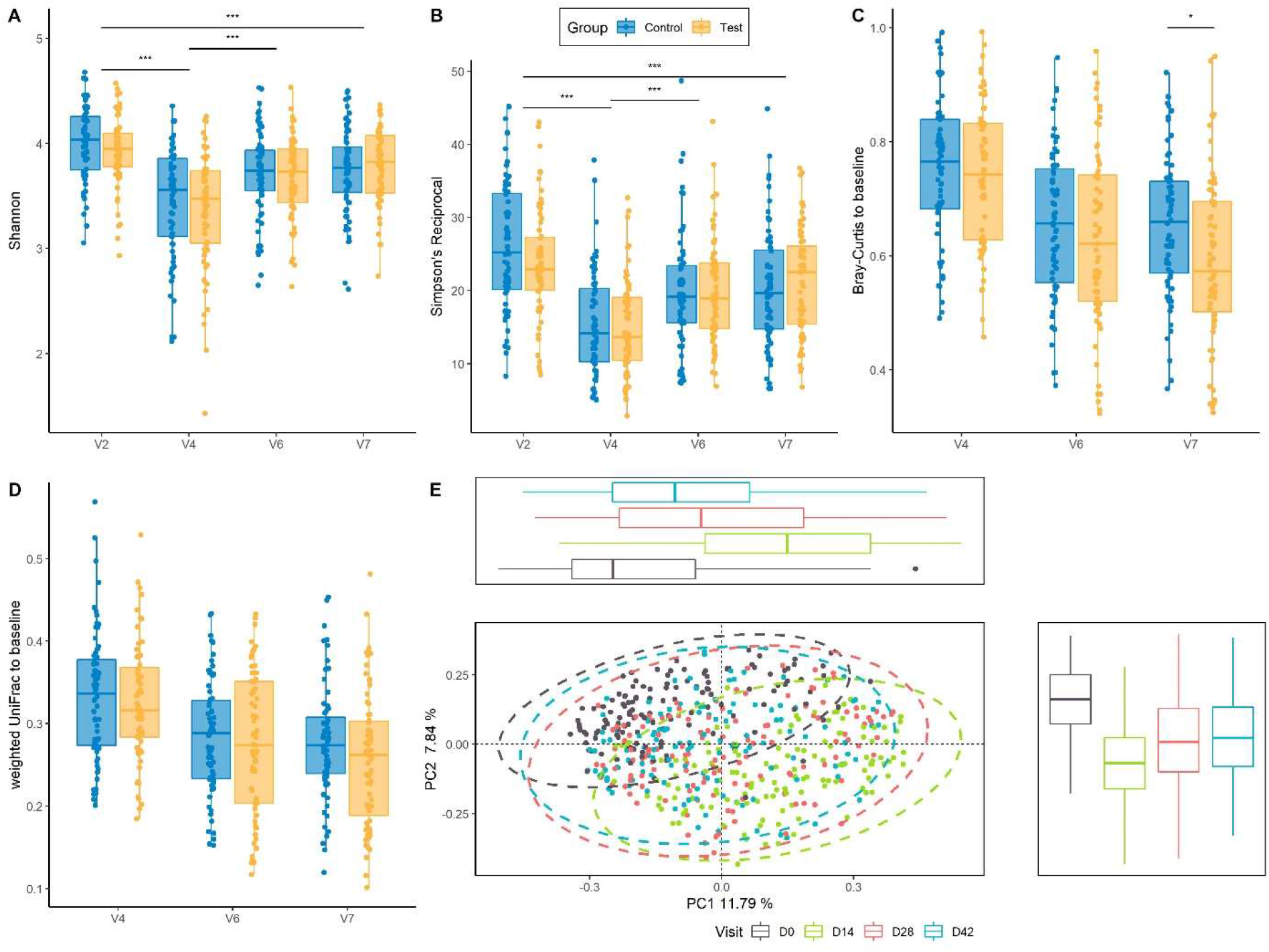
Figure S6. Global gut microbiota response to *Hp* treatment by Quantitative Microbiome Profiling. (A) Alpha-diversity assessed by Shannon index and (B) Alpha-diversity assessed by Simpson reciprocal (C) Intra-subject distance to baseline in Test and Control groups across the study based on Bray-Curtis dissimilarity (D) Intra-subject distance to baseline in Test and Control groups across the study based on Weighted UniFrac distance (E) Principal Coordinate analysis (PcoA) of Bray-Curtis dissimilarity. * p <0.05; ** p < 0.01; *** p < 0.001

**Additional file 14:**
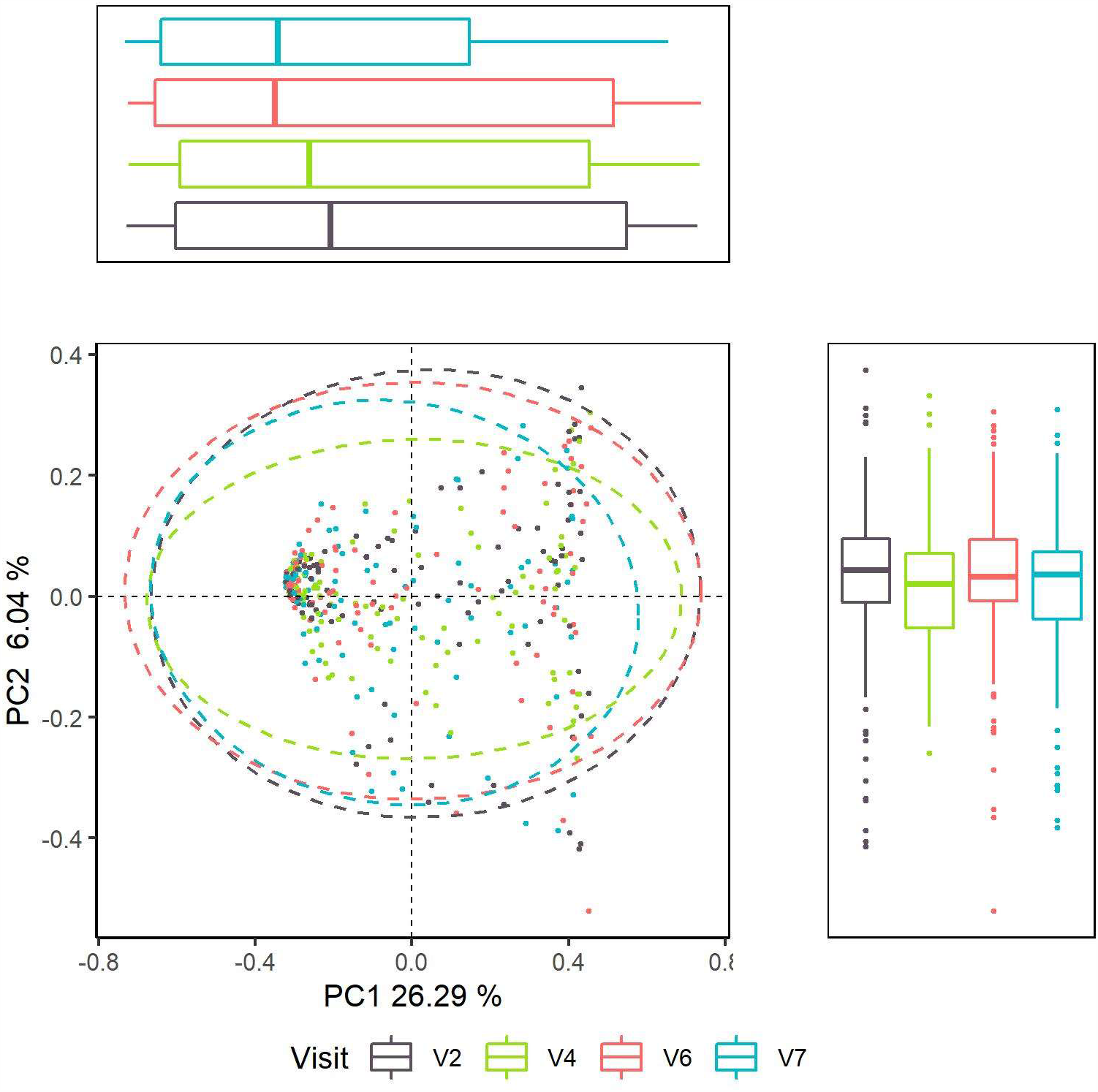
Figure S7. Global gut mycobiota response to *Hp* treatment. Principal Coordinate analysis (PcoA) of Bray-Curtis dissimilarity.

**Additional file 12:**
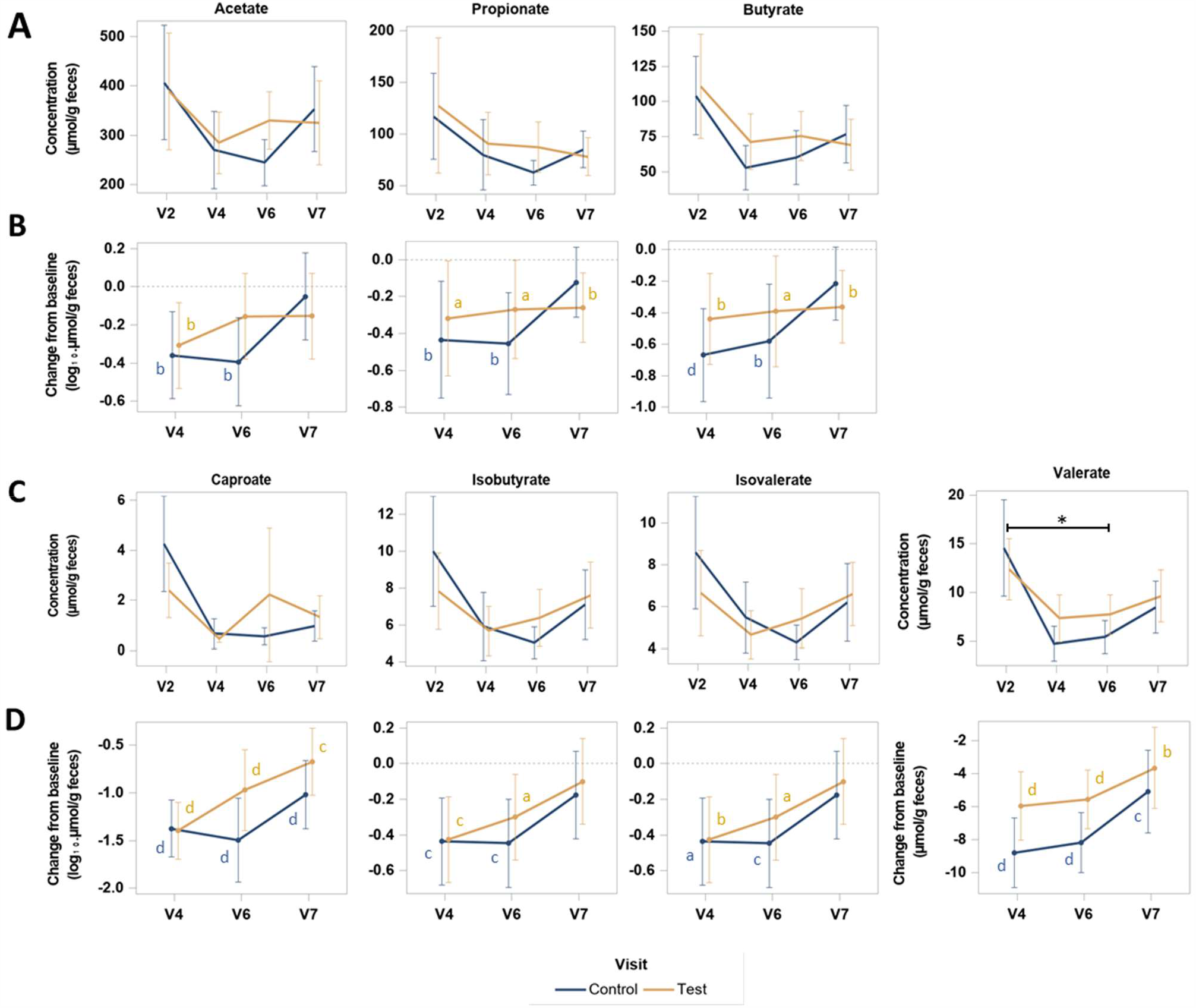
Figure S8. Quantification of individual fecal SCFA. Concentration (A, C) and change from baseline (B, D) of SCFA: acetate, propionate, butyrate (A, B), caproate, isobutyrate, isovalerate, valerate (C, D). P-values are provided according to Student test: * p <0.05 for comparison between groups of the change of valerate concentration from V2 to V6; ^a^ p <0.05, ^b^ p < 0.01; ^c^ p < 0.001; ^d^ p < 0.0001, for comparison within group of the change from V2 at each visit. SCFA concentrations are expressed in µmol/g of dry feces.

**Additional file 3:** Table S1. Bacterial strain counts in Test product during the shelf life.

| Strains | Shelf life | Mean (CFU/g)<sup>1</sup> | Min (CFU/g) | Max (CFU/g) |
| --- | --- | --- | --- | --- |
| *Lacticaseibacillus paracasei*<br>CNCM I-1518 | Day 3 | 2.85x10<sup>8</sup> | 1.50x10<sup>8</sup> | 5.00x10<sup>8</sup> |
|  | Day 42 | 1.85x10<sup>8</sup> | 1.20x10<sup>8</sup> | 3.90x10<sup>8</sup> |
| *Lacticaseibacillus paracasei*<br>CNCM I-3689 | Day 3 | 2.26x10<sup>7</sup> | 5.00x10<sup>6</sup> | 4.30x10<sup>7</sup> |
|  | Day 42 | 1.82x10<sup>7</sup> | 6.00x10<sup>6</sup> | 3.90x10<sup>7</sup> |
| *Lacticaseibacillus rhamnosus* CNCM I-3690 | Day 3 | 1.34x10<sup>8</sup> | 7.20x10<sup>7</sup> | 2.20x10<sup>8</sup> |
|  | Day 42 | 3.40x10<sup>7</sup> | 1.20x10<sup>7</sup> | 1.00x10<sup>8</sup> |
| *Streptococcus thermophilus*<br>(CNCM I-2773, CNCM I-2835, CNCM I-2778) | Day 3 | 5.56x10<sup>8</sup> | 3.70x10<sup>8</sup> | 7.50x10<sup>8</sup> |
|  | Day 42 | 5.44x10<sup>8</sup> | 2.60x10<sup>8</sup> | 8.10x10<sup>8</sup> |
| *Lactobacillus delbrueckii subsp bulgaricus*<br>CNCM I-2787 | Day 3 | 3.58x10<sup>6</sup> | 1.00x10<sup>5</sup> | 2.00x10<sup>7</sup> |
|  | Day 42 | 5.60x10<sup>2</sup> | 1.00x10<sup>1</sup> | 5.40x10<sup>3</sup> |

<sup>1</sup> : Means were calculated as derived from measures carried out in each of the 18 batches of product used in the study. Counts were measured for each of the bacterial strains present in the Test product, at the start (day 3) and end (day 42) of the authorized storage period (shelf life).

**Additional file 6:** Table S2. Number of days of GI symptoms.

|  |  | **Test (N=68)** | **Control (N=68)** |
| --- | --- | --- | --- |
| **Number of days with GI symptoms<sup>1</sup>** | Mean (SD) | 9.5 (8.0) | 8.4 (8.5) |
|  | Min ; Max | 0 ; 28 | 0 ; 28 |
| **Number of days with diarrhea<sup>2</sup>** | Mean (SD) | 0.2 (1.2) | 0.1 (0.5) |
|  | Min ; Max | 0 ; 8 | 0 ; 3 |
| **Number of days with abdominal pain** | Mean (SD) | 2.4 (3.4) | 2.4 (3.6) |
|  | Min ; Max | 0 ; 18 | 0 ; 13 |
| **Number of days bloating** | Mean (SD) | 8.1 (8.0) | 6.7 (8.1) |
|  | Min ; Max | 0 ; 28 | 0 ; 28 |
| **Number of days nausea** | Mean (SD) | 0.8 (1.4) | 1.4 (2.8) |
|  | Min ; Max | 0 ; 6 | 0 ; 14 |
| **Number of days vomiting** | Mean (SD) | 0.0 (0.1) | 0.1 (0.4) |
|  | Min ; Max | 0 ; 1 | 0 ; 2 |

<sup>1</sup>: Total cumulative number of days with GI symptoms between V2 and V6.
<sup>2</sup>: Based on AAD reporting according to AAD definition 2.

**Additional file 13:** Table S4. Adverse events.

|  | Test (N=68) |  | Control (N=68) |  | All (N=136) |  |
| --- | --- | --- | --- | --- | --- | --- |
|  | Event<sup>1</sup> | n (%)<sup>2</sup> | Event<sup>1</sup> | n (%)<sup>2</sup> | Event<sup>1</sup> | n (%)<sup>2</sup> |
| AEs | 91 | 42 (61.8) | 93 | 42 (61.8) | 184 | 84 (61.8) |
| Serious AEs | 0 | 0 | 1 | 1 (1.5) | 1 | 1 (0.7) |
| Severe AEs | 0 | 0 | 2 | 1 (1.5) | 2 | 1 (0.7) |
| AEs unlikely related to study product | 38 | 22 (32.4) | 55 | 30 (44.1) | 93 | 52 (38.2) |
| AEs causing study product interruption | 3 | 1 (1.5) | 0 | 0 | 3 | 1 (0.7) |
| AEs related to *Hp* eradication treatment | 53 | 28 (41.2) | 47 | 28 (41.2) | 100 | 56 (41.2) |
| AEs causing *Hp* treatment interruption | 7 | 4 (5.9) | 3 | 3 (4.4) | 10 | 7 (5.1) |

<sup>1</sup>: Number of AE. <sup>2</sup>: Number and percentage of subjects with AE. Subject or event in more than one category are counted in each of those categories. Related AE have been categorized as 'Unlikely', 'Possibly', 'Probably' or 'Definitely' related. Any events with a missing causality was assumed to be related.

